# Serological outcomes of SARS-CoV-2 infection by vaccination status and variant in England

**DOI:** 10.1101/2023.09.05.23295073

**Authors:** Catherine Quinot, Rachel Lunt, Freja Kirsebom, Catriona Skarnes, Nick Andrews, Heather Whitaker, Charlotte Gower, Louise Letley, Donna Haskins, Catriona Angel, Skye Firminger, Kay Ratcliffe, Angela Sherridan, Shelina Rajan, Lola Akindele, Samreen Ijaz, Maria Zambon, Kevin Brown, Mary Ramsay, Jamie Lopez Bernal

**Affiliations:** UK Health Security Agency, London, United Kingdom; NIHR Health Protection Research Unit in Vaccines and Immunisation, London School of Hygiene and Tropical Medicine, London, United Kingdom; NIHR Health Protection Research Unit in Respiratory Infections, Imperial College London, London, United Kingdom

**Keywords:** N antibody, S antibody, SARS-CoV-2, SARS-CoV-2 vaccination, SARS-CoV-2 variants

## Abstract

**Background:** Throughout the SARS-CoV-2 pandemic, several vaccines have been rolled out and distinct variants with different severity and immune profiles emerged in England. Using data from enhanced surveillance of COVID-19 in vaccine eligible individuals we investigated the antibody response following SARS-CoV-2 infection according to vaccination status and variant.

**Methods:** PCR-positive eligible individuals were identified from community PCR testing data in England between February 2021 and April 2022 and contacted by nurses to complete questionnaires at recruitment and 21 days post recruitment. Individuals were sent self-sampling kits and self-sampled nasal/oropharyngeal swabs were taken day 1, day 3 and day 7 post-recruitment as well as acute (day 1), convalescent (follow-up) serum and oral fluid samples. Regression analyses were used to investigate how N antibody seroconversion differs by vaccine status, and to investigate how N and S antibody levels differ by vaccine status overall and stratified by variants. Interval-censored analyses and regression analyses were used to investigate the effect of acute S antibody levels on the duration of positivity, the cycle threshold values, the self-reported symptom severity and the number of symptoms reported.

**Results:** A total of 1,497 PCR positive individuals were included. A total of 369 (24.7%) individuals were unvaccinated, 359 (24.0%) participants were infected with Alpha, 762 (50.9%) with Delta and 376 (25.2%) with Omicron. The median age of participants was 49 years old (IQR 39–57). Convalescent anti-N antibody levels were lower in vaccinated individuals and convalescent anti-S antibody levels were higher in vaccinated individuals and increased with the number of doses received. Acute anti-S antibody level increased with the number of doses received. Higher acute anti-S antibody levels were associated with a shorter duration of positivity (overall and for the Delta variant). Higher acute anti-S antibody levels were also associated with higher Ct values (overall and for the Alpha and Delta variants). There was no association between the acute anti-S antibody level and self-reported symptom severity. Individuals with higher acute anti-S antibody level were less likely to report six or more symptoms (overall and for Delta variant).

**Conclusion:** Understanding the characteristics of the antibody response, its dynamics over time and the immunity it confers is important to inform future vaccination strategies and policies. Our findings suggest that vaccination is associated with high acute anti-S antibody level but reduced convalescent anti-N antibody level. High anti-S antibody level is associated with reduced duration of infection, reduced infectiousness and may also be associated with reduced symptoms severity and number of symptoms.

## Introduction

Throughout the SARS-CoV-2 pandemic, several vaccines have been rolled out and distinct variants with different severity and immune profiles emerged in England (1). Alpha was the main variant before May 2021, Delta was dominant between May and November 2021 (2) and was more transmissible and severe (3). Omicron was dominant after December 2021 (2) and caused less severe disease than Delta (4).

Roll out of the COVID-19 immunisation programme began in England in December 2020. Vaccines were initially administered to priority groups, including care home residents, people ≥80 years old, healthcare workers and those clinically vulnerable (≥16 years), and then offered to the rest of the adult (≥18 years) population in decreasing age order (5). Both messenger ribonucleic acid (mRNA) vaccines (Pfizer BioNTech and Moderna) and adenoviral vector vaccines (AstraZeneca) were used for primary courses, but booster doses were primarily restricted to mRNA vaccines (Pfizer BioNTech and Moderna). Following the introduction of the immunisation programme, an enhanced surveillance system (ES) was set up by the UK Health Security Agency (UKHSA) in February 2021 to monitor COVID-19 in vaccinated individuals in England. One of the objectives of the enhanced surveillance system was to investigate the antibody response following SARS-CoV-2 infection according to vaccination status and variant.

SARS-CoV-2 has four main proteins; spike (S), envelope (E), membrane (M), and nucleocapsid (N) protein, serological assays are generally based on S and N (6). Given that all vaccines used in the UK are based on the spike protein, antibodies to nucleocapsid antigen (anti-N) are generated in response to natural infection whereas antibodies to spike antigen (anti-S) are generated in response to both natural infection and vaccination (7). Most of the evidence on the antibody response post infection is focused on specific occupational populations (e.g., health and social case professionals) (8–11). Although antibody levels wane following an initial steep rise after SARS-CoV-2 infection, immune memory persists for months and likely years (12). Individuals who naturally contracted SARS-CoV-2 are thought to develop a more rapid and sustained response to COVID-19 vaccines than individuals not previously infected (13).

There have been numerous investigations of vaccine effectiveness and immune response to infection. Studies globally demonstrated that the effectiveness of SARS-CoV-2 vaccines wane over time, but the total effect of anti-S antibody levels on the risk of becoming infected by SARS-CoV-2 and whether this varies by vaccine type is not well understood (14). Given the continuous evolution of SARS-CoV-2 and waning immunity, understanding the antibody response to vaccines and variants, and how this has evolved over time during the pandemic, is important to inform future vaccination strategies and policies. Furthermore, this may help to inform predictive vaccinology considerations in designing the most effective vaccines for the future.

The aims of this paper were to investigate the serological outcomes of SARS-CoV-2 infection among vaccinated and unvaccinated individuals during the Alpha, Delta and Omicron waves using prospective data collected as part of the ES of COVID-19 vaccines in England. The specific aims were to a) compare the proportion of individuals who seroconverted for anti-N antibody following infection by vaccination status and variant, b) compare the levels of convalescent anti-N and anti-S antibody by vaccination status and following infection with either Alpha, Delta or the Omicron variants, c) to investigate the anti-S antibody levels in acute sera by vaccination type, d) to investigate the effect of acute anti-S antibody levels on the subsequent disease course (duration of positivity, Ct values, self-reported symptom severity and number of symptoms).

## Methods

### Study design

Individuals were eligible if they were polymerase chain reaction (PCR)-positive for COVID-19, 18 years and older, English residents and vaccine-eligible at time of recruitment. Care home residents, individuals with a previous positive PCR test within 90 days of their more recent positive test or tested positive more than 10 days after symptoms onset or enrolled in the SARS-CoV-2 immunity and reinfection evaluation (SIREN) study were excluded (15).

Individuals were contacted by UKHSA nurses between February 2021 and April 2022 to complete questionnaires at recruitment and 21 days post recruitment. Self-sampled nasal/oropharyngeal swabs were taken day 1, day 3 and day 7 post-recruitment as well as acute (day 1), convalescent (follow-up) serum and oral fluid samples (Figure 1). All samples were tested at the national virus reference department (VRD) at UKHSA.

**Figure 1.**
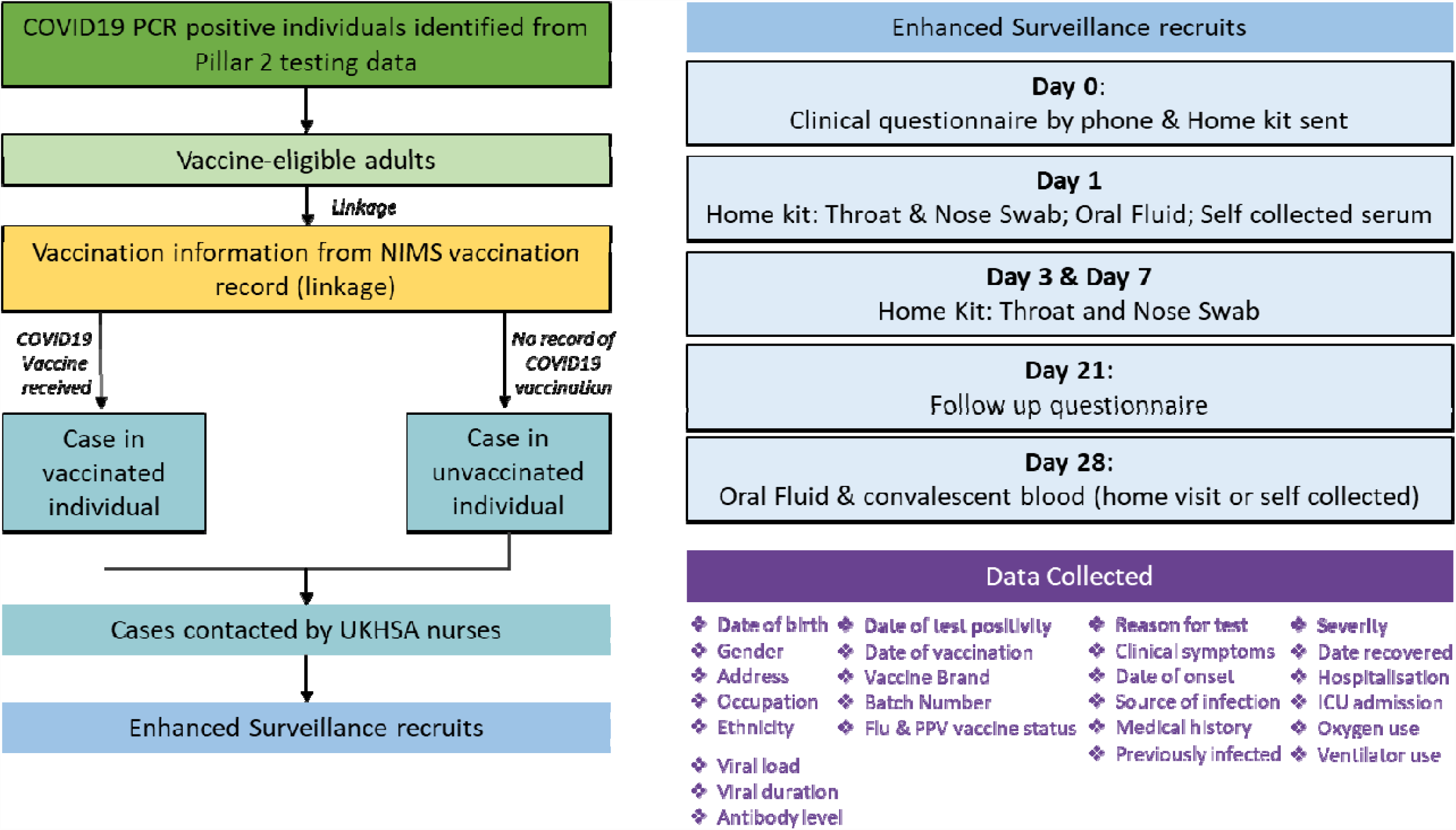
Flow chart summary of the Enhanced surveillance study of COVID-19 vaccines in England, 2021-2022.

### Data sources

Cases were recruited from community PCR testing (Pillar 2) data and linked to the National Immunisation Management System (NIMS)(16), an electronic register of all COVID-19 vaccinations given in England. Nurses collected information on demographics, symptoms, symptoms onset date, symptoms type, COVID-19 illness, severity, exposure risk factors, comorbidities at recruitment and 21 days post recruitment (Supplementary Figure 3). Serum samples were tested for the presence of antibodies to SARS-CoV-2 nucleoprotein (anti-N) and Spike Receptor Binding Domain (anti-S) using the “Elecsys anti-SARS-CoV-2 N” and the “Elecsys anti-SARS-CoV-2 S” electrochemiluminescence immunoassays respectively, on the Roche Cobas Pro e801 analyser. Oral fluid samples were tested for SARS-COV-2 antibodies against an N gene and an S gene target (17, 18). PCR cycle threshold (Ct) values were used as a semiquantitative measure of SARS-CoV-2 viral load (19). Variant status was confirmed by whole-genome sequencing of positive confirmatory PCR swabs for a subset of the samples, or sequencing results from original positive swab (where available). Details on the data sources are available in Supplementary Methods.

### Data selection

Individuals with a positive anti-N antibody result at recruitment (indicating the presence of naturally acquired COVID-19 antibodies), or who reported to have had a previous positive PCR or antibody test at recruitment, were excluded. Individuals with an original positive PCR test but no confirmatory positive follow up swab and a negative convalescent anti-N antibody result (suggesting no infection) were considered as false positive on their original test and excluded. Only individuals with qualitative result for whether an acute or a convalescent antibody result were included. Where an antibody result was missing, qualitative oral fluid results were used as a substitute. (Supplementary Figure 2).

### Definition of variables

Vaccination status was defined as unvaccinated, ≥28 days post dose 1, ≥14 days post dose 2, and ≥14 days post dose 3 or post dose 4. Vaccination status was then stratified according to vaccine type (mRNA vs adenovirus vector) (regardless of primary course). Vaccination date, dose and manufacturer from NIMS at the event date were used. The event date used throughout was the symptom onset date or the date of the original test where symptom start date was not available.

A blood sample was defined as acute for anti-N antibody if the interval between the sample date and the event date was less than 10 days (the N antibody response typically takes 10 days to get started). A blood sample was defined as acute for anti-S antibody if the interval between the sample date and the symptom onset date was less than 6 days (S antibodies in vaccinated individuals start to increase very quickly post infection by around 4 or 5 days). A blood sample was defined to be convalescent for anti-N and anti-S antibodies if the interval between the sample date and the event date was more than 21 days. For the analysis investigating the effect of acute anti-S antibody levels on the subsequent disease course, the antibody level was classified as 1: <1000 au/ml, 2: 1000-9999 au/ml and 3: ≥10,000 au/ml.

Comorbidities were self-reported (see questionnaire, Supplementary Figure 3). The time since event was defined as the interval between the event date and convalescent blood sample date.

### Statistical analysis

Anti-N seroconversion status (seroconversion of anti-N from negative to positive) by vaccination status and variant was investigated. Multivariable logistic regression was used with adjustment on self-reported gender, age, ethnicity, presence of comorbidities, immunosuppressed and clinically extremely vulnerable (CEV) status and time since event.

Convalescent anti-N and anti-S antibody geometric levels were estimated by vaccination status and variant. Multivariable linear regression on logged anti-N and anti-S antibody levels was used with adjustment on the variables defined below. Interaction between vaccination status and variants has been tested. The ratio between the convalescent and baseline anti-N antibody levels has been calculated to describe the change of antibody level by vaccination status and variants.

Anti-S antibody geometric mean levels in acute sera were estimated by vaccine type. Multivariable linear regression on logged anti-S antibody levels in acute sera was used with adjustment for self-reported gender, age, ethnicity, presence of comorbidities, immunosuppressed and CEV status and time since onset. The effect of anti-S antibody levels in acute sera on the disease in vaccinated individuals all variants combined and by variants was investigated. Multivariable logistic regression was performed to investigate the effect on the self-reported symptom severity. Interval censored survival analysis was used to investigate the effect on the duration of positivity (defined as the time from event date to the date of first negative test). Asymptomatic individuals were removed from symptom severity and duration of positivity analysis to avoid bias. Random effects linear regression was used to investigate the effect on the Ct values for both the ORF1ab and E genes of SARS-CoV-2 as a proxy for viral load. Only swabs with interpretable Ct values were included. Multivariable logistic regression was performed to investigate the effect on the number of symptoms (defined as 1-5 vs ≥6 symptoms). Models were adjusted on the variables defined previously.

## Results

### Description of the population

Of 1,973 eligible individuals, 60 (3%) were identified as a false positive and excluded (Supplementary Figure 2). A total of 1,497 individuals who had an original positive PCR result and no evidence of prior infection with SARS-CoV-2 (a positive anti-N antibody result at baseline or reporting to have a previous positive PCR or antibody test at recruitment suggested a prior infection) were included in the analysis. The characteristics of the population by vaccination status and variant are described in Table 1. A total of 369 (24.7%) individuals were unvaccinated, 180 (12.0%) were ≥28 days post dose 1, 608 (40.6%) were ≥14 days post dose 2, 340 (22.7%) were ≥14 days post dose 3 or post dose 4. Overall, 359 (24.0%) participants were infected with Alpha, 762 (50.9%) with Delta and 376 (25.2%) with Omicron. The median age of participants was 49 years old (IQR 39– 57, range 20-88 years old). Participants were spread throughout the country and were mainly of white ethnicity and with a female to male ratio of 3:2. Nearly half reported to have comorbidities with the most common being asthma and hypertension.

**Table 1.**
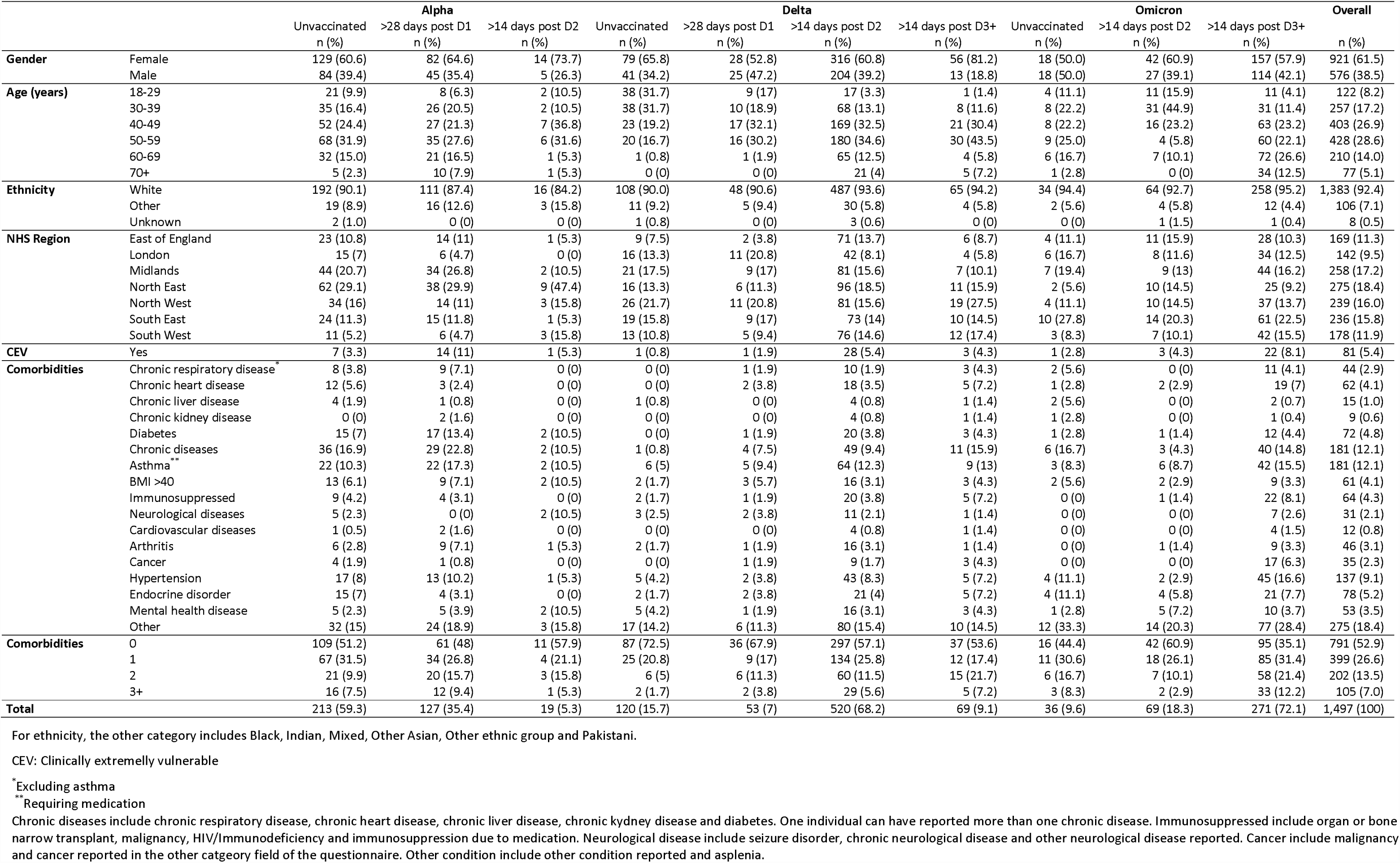
Characteristics of the individuals included in the analysis by vaccination status and variants in England, N=1,497.

|  |  | Alpha |  |  | Delta |  |  |  | Omicron |  |  | Overall |
| --- | --- | --- | --- | --- | --- | --- | --- | --- | --- | --- | --- | --- |
|  |  | Unvaccinated<br>n (%) | >28 days post D1<br>n (%) | >14 days post D2<br>n (%) | Unvaccinated<br>n (%) | >28 days post D1<br>n (%) | >14 days post D2<br>n (%) | >14 days post D3+<br>n (%) | Unvaccinated<br>n (%) | >14 days post D2<br>n (%) | >14 days post D3+<br>n (%) | n (%) |
| Gender | Female | 129 (60.6) | 82 (64.6) | 14 (73.7) | 79 (65.8) | 28 (52.8) | 316 (60.8) | 56 (81.2) | 18 (50.0) | 42 (60.9) | 157 (57.9) | 921 (61.5) |
|  | Male | 84 (39.4) | 45 (35.4) | 5 (26.3) | 41 (34.2) | 25 (47.2) | 204 (39.2) | 13 (18.8) | 18 (50.0) | 27 (39.1) | 114 (42.1) | 576 (38.5) |
| Age (years) | 18-29 | 21 (9.9) | 8 (6.3) | 2 (10.5) | 38 (31.7) | 9 (17) | 17 (3.3) | 1 (1.4) | 4 (11.1) | 11 (15.9) | 11 (4.1) | 122 (8.2) |
|  | 30-39 | 35 (16.4) | 26 (20.5) | 2 (10.5) | 38 (31.7) | 10 (18.9) | 68 (13.1) | 8 (11.6) | 8 (22.2) | 31 (44.9) | 31 (11.4) | 257 (17.2) |
|  | 40-49 | 52 (24.4) | 27 (21.3) | 7 (36.8) | 23 (19.2) | 17 (32.1) | 169 (32.5) | 21 (30.4) | 8 (22.2) | 16 (23.2) | 63 (23.2) | 403 (26.9) |
|  | 50-59 | 68 (31.9) | 35 (27.6) | 6 (31.6) | 20 (16.7) | 16 (30.2) | 180 (34.6) | 30 (43.5) | 9 (25.0) | 4 (5.8) | 60 (22.1) | 428 (28.6) |
|  | 60-69 | 32 (15.0) | 21 (16.5) | 1 (5.3) | 1 (0.8) | 1 (1.9) | 65 (12.5) | 4 (5.8) | 6 (16.7) | 7 (10.1) | 72 (26.6) | 210 (14.0) |
|  | 70+ | 5 (2.3) | 10 (7.9) | 1 (5.3) | 0 (0) | 0 (0) | 21 (4) | 5 (7.2) | 1 (2.8) | 0 (0) | 34 (12.5) | 77 (5.1) |
| Ethnicity | White | 192 (90.1) | 111 (87.4) | 16 (84.2) | 108 (90.0) | 48 (90.6) | 487 (93.6) | 65 (94.2) | 34 (94.4) | 64 (92.7) | 258 (95.2) | 1,383 (92.4) |
|  | Other | 19 (8.9) | 16 (12.6) | 3 (15.8) | 11 (9.2) | 5 (9.4) | 30 (5.8) | 4 (5.8) | 2 (5.6) | 4 (5.8) | 12 (4.4) | 106 (7.1) |
|  | Unknown | 2 (1.0) | 0 (0) | 0 (0) | 1 (0.8) | 0 (0) | 3 (0.6) | 0 (0) | 0 (0) | 1 (1.5) | 1 (0.4) | 8 (0.5) |
| NHS Region | East of England | 23 (10.8) | 14 (11) | 1 (5.3) | 9 (7.5) | 2 (3.8) | 71 (13.7) | 6 (8.7) | 4 (11.1) | 11 (15.9) | 28 (10.3) | 169 (11.3) |
|  | London | 15 (7) | 6 (4.7) | 0 (0) | 16 (13.3) | 11 (20.8) | 42 (8.1) | 4 (5.8) | 6 (16.7) | 8 (11.6) | 34 (12.5) | 142 (9.5) |
|  | Midlands | 44 (20.7) | 34 (26.8) | 2 (10.5) | 21 (17.5) | 9 (17) | 81 (15.6) | 7 (10.1) | 7 (19.4) | 9 (13) | 44 (16.2) | 258 (17.2) |
|  | North East | 62 (29.1) | 38 (29.9) | 9 (47.4) | 16 (13.3) | 6 (11.3) | 96 (18.5) | 11 (15.9) | 2 (5.6) | 10 (14.5) | 25 (9.2) | 275 (18.4) |
|  | North West | 34 (16) | 14 (11) | 3 (15.8) | 26 (21.7) | 11 (20.8) | 81 (15.6) | 19 (27.5) | 4 (11.1) | 10 (14.5) | 37 (13.7) | 239 (16.0) |
|  | South East | 24 (11.3) | 15 (11.8) | 1 (5.3) | 19 (15.8) | 9 (17) | 73 (14) | 10 (14.5) | 10 (27.8) | 14 (20.3) | 61 (22.5) | 236 (15.8) |
|  | South West | 11 (5.2) | 6 (4.7) | 3 (15.8) | 13 (10.8) | 5 (9.4) | 76 (14.6) | 12 (17.4) | 3 (8.3) | 7 (10.1) | 42 (15.5) | 178 (11.9) |
| CEV | Yes | 7 (3.3) | 14 (11) | 1 (5.3) | 1 (0.8) | 1 (1.9) | 28 (5.4) | 3 (4.3) | 1 (2.8) | 3 (4.3) | 22 (8.1) | 81 (5.4) |
| Comorbidities | Chronic respiratory disease* | 8 (3.8) | 9 (7.1) | 0 (0) | 0 (0) | 1 (1.9) | 10 (1.9) | 3 (4.3) | 2 (5.6) | 0 (0) | 11 (4.1) | 44 (2.9) |
|  | Chronic heart disease | 12 (5.6) | 3 (2.4) | 0 (0) | 0 (0) | 2 (3.8) | 18 (3.5) | 5 (7.2) | 1 (2.8) | 2 (2.9) | 19 (7) | 62 (4.1) |
|  | Chronic liver disease | 4 (1.9) | 1 (0.8) | 0 (0) | 1 (0.8) | 0 (0) | 4 (0.8) | 1 (1.4) | 2 (5.6) | 0 (0) | 2 (0.7) | 15 (1.0) |
|  | Chronic kidney disease | 0 (0) | 2 (1.6) | 0 (0) | 0 (0) | 0 (0) | 4 (0.8) | 1 (1.4) | 1 (2.8) | 0 (0) | 1 (0.4) | 9 (0.6) |
|  | Diabetes | 15 (7) | 17 (13.4) | 2 (10.5) | 0 (0) | 1 (1.9) | 20 (3.8) | 3 (4.3) | 1 (2.8) | 1 (1.4) | 12 (4.4) | 72 (4.8) |
|  | Chronic diseases | 36 (16.9) | 29 (22.8) | 2 (10.5) | 1 (0.8) | 4 (7.5) | 49 (9.4) | 11 (15.9) | 6 (16.7) | 3 (4.3) | 40 (14.8) | 181 (12.1) |
|  | Asthma** | 22 (10.3) | 22 (17.3) | 2 (10.5) | 6 (5) | 5 (9.4) | 64 (12.3) | 9 (13) | 3 (8.3) | 6 (8.7) | 42 (15.5) | 181 (12.1) |
|  | BMI >40 | 13 (6.1) | 9 (7.1) | 2 (10.5) | 2 (1.7) | 3 (5.7) | 16 (3.1) | 3 (4.3) | 2 (5.6) | 2 (2.9) | 9 (3.3) | 61 (4.1) |
|  | Immunosuppressed | 9 (4.2) | 4 (3.1) | 0 (0) | 2 (1.7) | 1 (1.9) | 20 (3.8) | 5 (7.2) | 0 (0) | 1 (1.4) | 22 (8.1) | 64 (4.3) |
|  | Neurological diseases | 5 (2.3) | 0 (0) | 2 (10.5) | 3 (2.5) | 2 (3.8) | 11 (2.1) | 1 (1.4) | 0 (0) | 0 (0) | 7 (2.6) | 31 (2.1) |
|  | Cardiovascular diseases | 1 (0.5) | 2 (1.6) | 0 (0) | 0 (0) | 0 (0) | 4 (0.8) | 1 (1.4) | 0 (0) | 0 (0) | 4 (1.5) | 12 (0.8) |
|  | Arthritis | 6 (2.8) | 9 (7.1) | 1 (5.3) | 2 (1.7) | 1 (1.9) | 16 (3.1) | 1 (1.4) | 0 (0) | 1 (1.4) | 9 (3.3) | 46 (3.1) |
|  | Cancer | 4 (1.9) | 1 (0.8) | 0 (0) | 0 (0) | 1 (1.9) | 9 (1.7) | 3 (4.3) | 0 (0) | 0 (0) | 17 (6.3) | 35 (2.3) |
|  | Hypertension | 17 (8) | 13 (10.2) | 1 (5.3) | 5 (4.2) | 2 (3.8) | 43 (8.3) | 5 (7.2) | 4 (11.1) | 2 (2.9) | 45 (16.6) | 137 (9.1) |
|  | Endocrine disorder | 15 (7) | 4 (3.1) | 0 (0) | 2 (1.7) | 2 (3.8) | 21 (4) | 5 (7.2) | 4 (11.1) | 4 (5.8) | 21 (7.7) | 78 (5.2) |
|  | Mental health disease | 5 (2.3) | 5 (3.9) | 2 (10.5) | 5 (4.2) | 1 (1.9) | 16 (3.1) | 3 (4.3) | 1 (2.8) | 5 (7.2) | 10 (3.7) | 53 (3.5) |
|  | Other | 32 (15) | 24 (18.9) | 3 (15.8) | 17 (14.2) | 6 (11.3) | 80 (15.4) | 10 (14.5) | 12 (33.3) | 14 (20.3) | 77 (28.4) | 275 (18.4) |
| Comorbidities | 0 | 109 (51.2) | 61 (48) | 11 (57.9) | 87 (72.5) | 36 (67.9) | 297 (57.1) | 37 (53.6) | 16 (44.4) | 42 (60.9) | 95 (35.1) | 791 (52.9) |
|  | 1 | 67 (31.5) | 34 (26.8) | 4 (21.1) | 25 (20.8) | 9 (17) | 134 (25.8) | 12 (17.4) | 11 (30.6) | 18 (26.1) | 85 (31.4) | 399 (26.6) |
|  | 2 | 21 (9.9) | 20 (15.7) | 3 (15.8) | 6 (5) | 6 (11.3) | 60 (11.5) | 15 (21.7) | 6 (16.7) | 7 (10.1) | 58 (21.4) | 202 (13.5) |
|  | 3+ | 16 (7.5) | 12 (9.4) | 1 (5.3) | 2 (1.7) | 2 (3.8) | 29 (5.6) | 5 (7.2) | 3 (8.3) | 2 (2.9) | 33 (12.2) | 105 (7.0) |
| Total |  | 213 (59.3) | 127 (35.4) | 19 (5.3) | 120 (15.7) | 53 (7) | 520 (68.2) | 69 (9.1) | 36 (9.6) | 69 (18.3) | 271 (72.1) | 1,497 (100) |

For ethnicity, the other category includes Black, Indian, Mixed, Other Asian, Other ethnic group and Pakistani.
CEV: Clinically extremely vulnerable
\*Excluding asthma
\*\*Requiring medication
Chronic diseases include chronic respiratory disease, chronic heart disease, chronic liver disease, chronic kidney disease and diabetes. One individual can have reported more than one chronic disease. Immunosuppressed include organ or bone marrow transplant, malignancy, HIV/immunodeficiency and immunosuppression due to medication. Neurological disease include seizure disorder, chronic neurological disease and other neurological disease reported. Cancer include malignancy and cancer reported in the other category field of the questionnaire. Other condition include other condition reported and asplenia.

A total of 1,464 (97.8%) individuals reported to be symptomatic and were included in the analysis of symptom severity and the duration of positivity. The characteristics of these individuals are described in Supplementary Table 1.

### Description of the serum and oral fluid samples

The characteristics of the acute and convalescent serum and oral fluid samples are shown in Supplementary Table 2 and Table 3. The median time from symptom onset date or original PCR test date to sample collection for the acute serum samples was 6 days (IQR: 5-7 days, range 3–13 days), however only samples up to five days post onset were used in analyses of anti-S antibodies in acute sera. Convalescent serum samples were collected on average 39 days (IQR: 35-44 days, range 22–88 days) after symptom onset date or original PCR test date.

#### Anti-N antibody seroconversion

Out of 1,115 individuals with an acute and convalescent serum sample or oral fluid sample with an anti-N antibody result, 1,065 seroconverted (95.5%). The description of the proportion of anti-N antibody seroconversion by vaccination status and variant is described in Supplementary Table 4.

Among the individuals who seroconverted, the geometric mean of the interval between the date of the original test/symptom onset and the sample date for the final blood was 40 days (min-max:22-88 days). Most of the seroconverted individuals returned their second blood/oral fluid sample between 35 and 41 days after the original test/symptom onset date (43.0%), 0.7% returned it before 28 days, 21.4% between 28 and 34 days, 21.8% between 42 and 48 days and 13.1% at 49 days or after.

Of the 50 participants that did not seroconvert, the median age was 49 years old, (IQR 39–58, range 22-81 years old), 76% were female, 98% were of white ethnicity, 8% were immunosuppressed, 12% were CEV, 16% reported to have three or more comorbidities, 22% reported to have chronic diseases and 10% reported to have chronic heart disease (Supplementary Table 5). The geometric mean (95% CI) of the convalescent anti-N antibody for these individuals was 0.34 (0.27-042), (min-max: 0.07-0.98).

After adjustment for age, gender, ethnicity, time since event, variant, immunosuppression and CEV status, and presence of comorbidities, no statistically significant association was found between the lack of anti-N antibody seroconversion and the vaccination status (Table 2). There was no significant difference between the lack of anti-N antibody seroconversion and variants, gender, ethnicity, age, comorbidity and immunosuppressed and CEV status (Table 2).

**Table 2.** Multivariable logistic regression investigating the variables associated to the lack of anti-N seroconversion.

|  | n=1,073 | Odds Ratio [95% CI] |
| --- | --- | --- |
| **Vaccination status** |  |  |
| Unvaccinated | 263 | 1.35 [0.51-3.57] |
| >28 days post D1 | 147 | 1.35 [0.43-4.25] |
| >14 days post D2 | 442 | 1 |
| >14 days post D3+ | 221 | 1.48 [0.53-4.11] |
| **Variants** |  |  |
| Alpha | 280 | 1.72 [0.68-4.41] |
| Delta | 556 | 1 |
| Omicron | 237 | 1.60 [0.60-4.27] |
| **Gender** |  |  |
| Female | 679 | 1 |
| Male | 394 | 0.55 [0.28-1.09] |
| **Ethnicity** |  |  |
| White | 1,004 | 1 |
| Other | 69 | 0.28 [0.04-2.09] |
| **Age (years old)** |  |  |
| 18-29 | 85 | 0.83 [0.26-2.65] |
| 30-39 | 187 | 0.90 [0.38-2.16] |
| 40-49 | 292 | 0.78 [0.36-1.70] |
| 50-59 | 309 | 1 |
| 60-69 | 151 | 0.66 [0.24-1.83] |
| 70+ | 49 | 0.71 [0.15-3.38] |
| **Comorbidity** |  |  |
| No | 574 | 1 |
| Yes | 499 | 0.70 [0.36-1.38] |
| **Immunosuppressed and CEV status** |  |  |
| Neither | 992 | 1 |
| Immunosuppressed only | 21 | 1.25 [0.15-10.28] |
| Immunosuppressed and/or CEV | 22 | 3.78 [0.98-14.66] |
| CEV only | 38 | 2.45 [0.65-9.23] |
| **Time since symptom onset (days)** | 1,073 | 0.99 [0.95-1.03] |

#### Convalescent anti-N antibody levels by vaccination status

Vaccinated individuals had significantly lower convalescent anti-N antibody levels compare to unvaccinated individuals (Table 3), this held true when analyses were stratified by variant. Immunosuppressed and CEV individuals generally had lower convalescent anti-N antibody levels than individuals not immunosuppressed and not CEV, this held true for Delta when stratified by variants (Supplementary Table 6).

**Table 3.** Multivariable linear regression investigating the convalescent anti-N antibody level by vaccination status for all variants and by variants - geometric mean (95% CI) and adjusted geometric mean ratio (95% CI) of the log of convalescent Roche N are presented.

|  | n | Geometric mean (95% CI) | Adjusted geometric mean ratio (95% CI) |
| --- | --- | --- | --- |
| **All variants** | 1,214 |  |  |
| Unvaccinated | 296 | 24.6 (20.5-29.4) | 1 |
| >28 days post D1 | 156 | 11.8 (9.3-15.1) | 0.46 (0.34-0.60)* |
| >14 days post D2 | 511 | 13.1 (11.6-14.8) | 0.41 (0.32-0.53)* |
| >14 days post D3+ | 251 | 11.7 (9.9-13.9) | 0.48 (0.34-0.67)* |
| **Alpha** | 304 |  |  |
| Unvaccinated | 176 | 22.6 (17.7-28.8) | 1 |
| >28 days post D1 | 110 | 10.4 (7.7-14.0) | 0.40 (0.28-0.59)* |
| >14 days post D2 | 18 | 10.5 (4.9-22.3) | 0.47 (0.22-1.00) |
| **Delta** | 644 |  |  |
| Unvaccinated | 96 | 30.3 (22.3-41.0) | 1 |
| >28 days post D1 | 46 | 16.1 (10.6-24.4) | 0.48 (0.29-0.79)* |
| >14 days post D2 | 447 | 13.8 (12.2-15.8) | 0.40 (0.28-0.56)* |
| >14 days post D3+ | 55 | 12.9 (8.9-18.7) | 0.40 (0.25-0.65)* |
| **Omicron** | 262 |  |  |
| Unvaccinated | 24 | 19.6 (11.5-33.4) | 1 |
| >14 days post D2 | 46 | 8.4 (5.4-13.1) | 0.46 (0.23-0.94)* |
| >14 days post D3+ | 196 | 11.4 (9.4-13.9) | 0.62 (0.33-1.15) |

Adjusted for age, gender, ethnicity, time since event, presence of comorbidities, immunosuppressed and clinically extremely vulnerable status
\* p<0.05

Convalescent anti-N antibody levels significantly differed by gender, age and ethnicity. Male, and other ethnic groups had higher convalescent anti-N antibody levels compared to female and white ethnic group. The convalescent anti-N antibody level did not significantly differ by presence of comorbidities. When stratified by variant, for Delta, other ethnic groups had higher convalescent anti-N antibody levels compared to white ethnic group. For Alpha, Delta and Omicron levels did not significantly differ by age, gender, and presence of comorbidities (Supplementary Table 6).

When looking at the ratio of convalescent and baseline anti-N antibody levels, a bigger ratio was found for the unvaccinated vs vaccinated individuals overall and when stratified by variant (Supplementary Table 7).

#### Anti-S antibody level in acute sera by vaccine type in vaccinated individuals

Overall, the adjusted geometric mean ratios were not statistically significant for all vaccine types and doses but generally the anti-S antibody levels in acute sera tended to be greater with increasing numbers of doses and tended to be greater for mRNA vaccines than adenoviral vector vaccine. Anti-S antibody levels in acute sera did not differ by age, gender, ethnicity, immunosuppressed and/or CEV status and presence of comorbidities (Supplementary Table 9).

#### Convalescent anti-S antibody levels by vaccination status

The adjusted geometric mean ratios were not statistically significant for all vaccine types and doses. Convalescent anti-S antibody levels tended to be greater with increasing numbers of vaccine doses and tended to be greater for mRNA vaccines (Pfizer/BioNTech or Moderna) than adenoviral vector vaccine (AstraZeneca) vaccine (Table 4). This held true when analyses were stratified by variant.

**Table 4.**
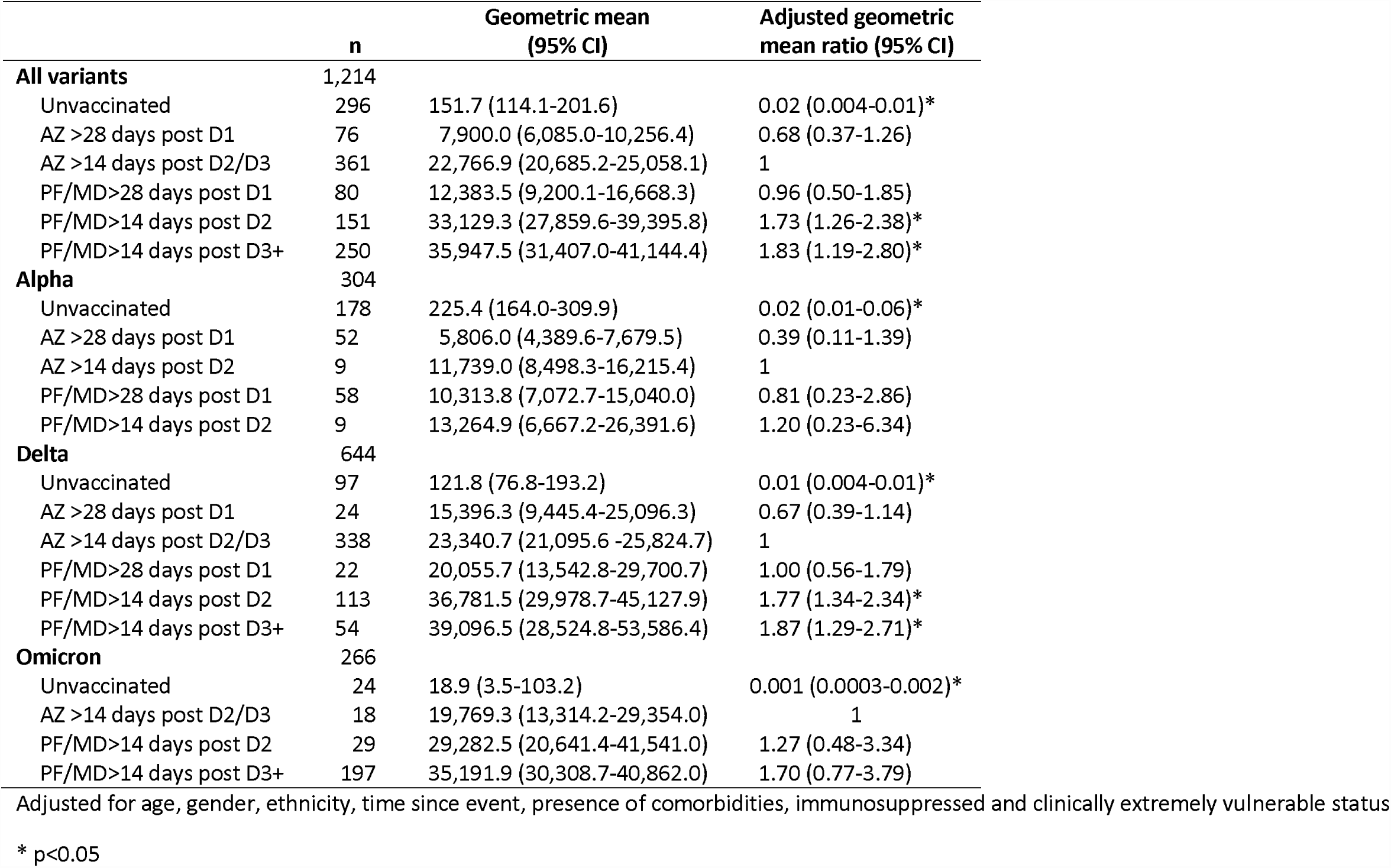
Multivariable linear regression investigating the convalescent anti-S antibody level by vaccination status for all variants combined and by variants - geometric mean (95% CI) and adjusted geometric mean ratio (95% CI) of the log of convalescent Roche S are presented.

|  | n | Geometric mean<br>(95% CI) | Adjusted geometric<br>mean ratio (95% CI) |
| --- | --- | --- | --- |
| **All variants** | 1,214 |  |  |
| Unvaccinated | 296 | 151.7 (114.1-201.6) | 0.02 (0.004-0.01)* |
| AZ >28 days post D1 | 76 | 7,900.0 (6,085.0-10,256.4) | 0.68 (0.37-1.26) |
| AZ >14 days post D2/D3 | 361 | 22,766.9 (20,685.2-25,058.1) | 1 |
| PF/MD>28 days post D1 | 80 | 12,383.5 (9,200.1-16,668.3) | 0.96 (0.50-1.85) |
| PF/MD>14 days post D2 | 151 | 33,129.3 (27,859.6-39,395.8) | 1.73 (1.26-2.38)* |
| PF/MD>14 days post D3+ | 250 | 35,947.5 (31,407.0-41,144.4) | 1.83 (1.19-2.80)* |
| **Alpha** | 304 |  |  |
| Unvaccinated | 178 | 225.4 (164.0-309.9) | 0.02 (0.01-0.06)* |
| AZ >28 days post D1 | 52 | 5,806.0 (4,389.6-7,679.5) | 0.39 (0.11-1.39) |
| AZ >14 days post D2 | 9 | 11,739.0 (8,498.3-16,215.4) | 1 |
| PF/MD>28 days post D1 | 58 | 10,313.8 (7,072.7-15,040.0) | 0.81 (0.23-2.86) |
| PF/MD>14 days post D2 | 9 | 13,264.9 (6,667.2-26,391.6) | 1.20 (0.23-6.34) |
| **Delta** | 644 |  |  |
| Unvaccinated | 97 | 121.8 (76.8-193.2) | 0.01 (0.004-0.01)* |
| AZ >28 days post D1 | 24 | 15,396.3 (9,445.4-25,096.3) | 0.67 (0.39-1.14) |
| AZ >14 days post D2/D3 | 338 | 23,340.7 (21,095.6 -25,824.7) | 1 |
| PF/MD>28 days post D1 | 22 | 20,055.7 (13,542.8-29,700.7) | 1.00 (0.56-1.79) |
| PF/MD>14 days post D2 | 113 | 36,781.5 (29,978.7-45,127.9) | 1.77 (1.34-2.34)* |
| PF/MD>14 days post D3+ | 54 | 39,096.5 (28,524.8-53,586.4) | 1.87 (1.29-2.71)* |
| **Omicron** | 266 |  |  |
| Unvaccinated | 24 | 18.9 (3.5-103.2) | 0.001 (0.0003-0.002)* |
| AZ >14 days post D2/D3 | 18 | 19,769.3 (13,314.2-29,354.0) | 1 |
| PF/MD>14 days post D2 | 29 | 29,282.5 (20,641.4-41,541.0) | 1.27 (0.48-3.34) |
| PF/MD>14 days post D3+ | 197 | 35,191.9 (30,308.7-40,862.0) | 1.70 (0.77-3.79) |

Adjusted for age, gender, ethnicity, time since event, presence of comorbidities, immunosuppressed and clinically extremely vulnerable status
\* p<0.05

Convalescent anti-S antibody level significantly differed by age but did not significantly differ by gender, ethnicity, immunosuppressed and CEV status and presence of comorbidities. When stratified by variant, convalescent anti-S antibody level did not significantly differ by age (except for Delta), gender, ethnicity, immunosuppressed and CEV status and presence of comorbidities (Supplementary Table 8).

#### Effect of anti-S antibody level in acute sera on the subsequent disease course

Individuals with a level of 10,000 au/ml or more acute anti-S antibody had significantly shorter duration of positivity compared to individuals with 1,000 to 9,999 au/ml antibody level overall (predicted medians (95%CI): 9.8 (8.4-11.3) days versus 12.3 (10.8-13.7) days respectively). When stratified by variant, there was a downwards trend in duration of positivity with increasing anti-S antibody for all variants The difference was significant for Delta (predicted medians (95%CI):10.0 (8.2-11.8) days versus 13.4 (10.6-16.2) days respectively) but not for Alpha or Omicron. (Table 5).

**Table 5.** a)Interval censored survival regression analysis assessing the effect of anti-S antibody level in acute sera on duration of positivity by all variants combined and by variants with b) predicted median time from positive to negative SARS-CoV-2 PCR test (duration of positivity)

|  | **All variants (n=110)** |  | **Alpha (n=18)** |  | **Delta (n=71)** |  | **Omicron (n=21)** |  |
| --- | --- | --- | --- | --- | --- | --- | --- | --- |
| **Acute S antibody level (au/ml)** | n | HR (95% CI) | n | HR (95% CI) | n | HR (95% CI) | n | HR (95% CI) |
| **<1,000** | 28 | 0.59 (0.25-1.43) | 9 | 0.001 (1.80e-06-1.77) | 19 | 0.58 (0.19-1.74) | 0 | N/A |
| **1,000-9,999** | 57 | 1 | 9 | 1 | 34 | 1 | 14 | 1 |
| **>=10,000** | 25 | 2.37 (1.11-5.07)* | 0 | N/A | 18 | 2.92 (1.20-7.13)* | 7 | 2.00 (0.35-11.49) |

|  | **All variants (n=110)** | **Alpha (n=18)** | **Delta (n=71)** | **Omicron (n=21)** |
| --- | --- | --- | --- | --- |
| **Acute S antibody level (au/ml)** | Median (95% CI) | Median (95% CI) | Median (95% CI) | Median (95% CI) |
| **<1,000** | 14.0 (11.2-16.8) | 18.6 (-3427.8-3465.0) | 15.5 (11.1-19.9) | N/A |
| **1,000-9,999** | 12.3 (10.8-13.7) | 11.4 (-2112.1-2134.9) | 13.4 (10.6-16.2) | 12.1 (10.0-14.1) |
| **>=10,000** | 9.8 (8.4-11.3) | N/A | 10.0 (8.2-11.8) | 10.9 (8.8-12.9) |

Individuals with a level of less than 1,000 au/ml acute anti-S antibody had significantly lower Ct values (higher viral load) compared to individuals with 1,000 to 9,999 au/ml antibody level overall (E gene Ct difference: −2.71, 95% CI: −4.20 to −1.22, Orf1ab gene Ct difference: −2.77, 95% CI: −4.33 to −1.21) When stratified by variant, there was an increasing trend in mean Ct values with increasing anti-S antibody for all variants. The difference was significant for Alpha (E gene Ct difference: −8.72, 95% CI: −15.14 to −2.31, Orf1ab gene Ct difference: −9.81, 95% CI: −16.00 to −3.63) and Delta (E gene Ct difference: −2.41, 95% CI: −4.04 to −0.77, Orf1ab gene Ct difference: −2.66, 95% CI: −4.42 to −0.89) but not for Omicron (Table 6).

**Table 6.**
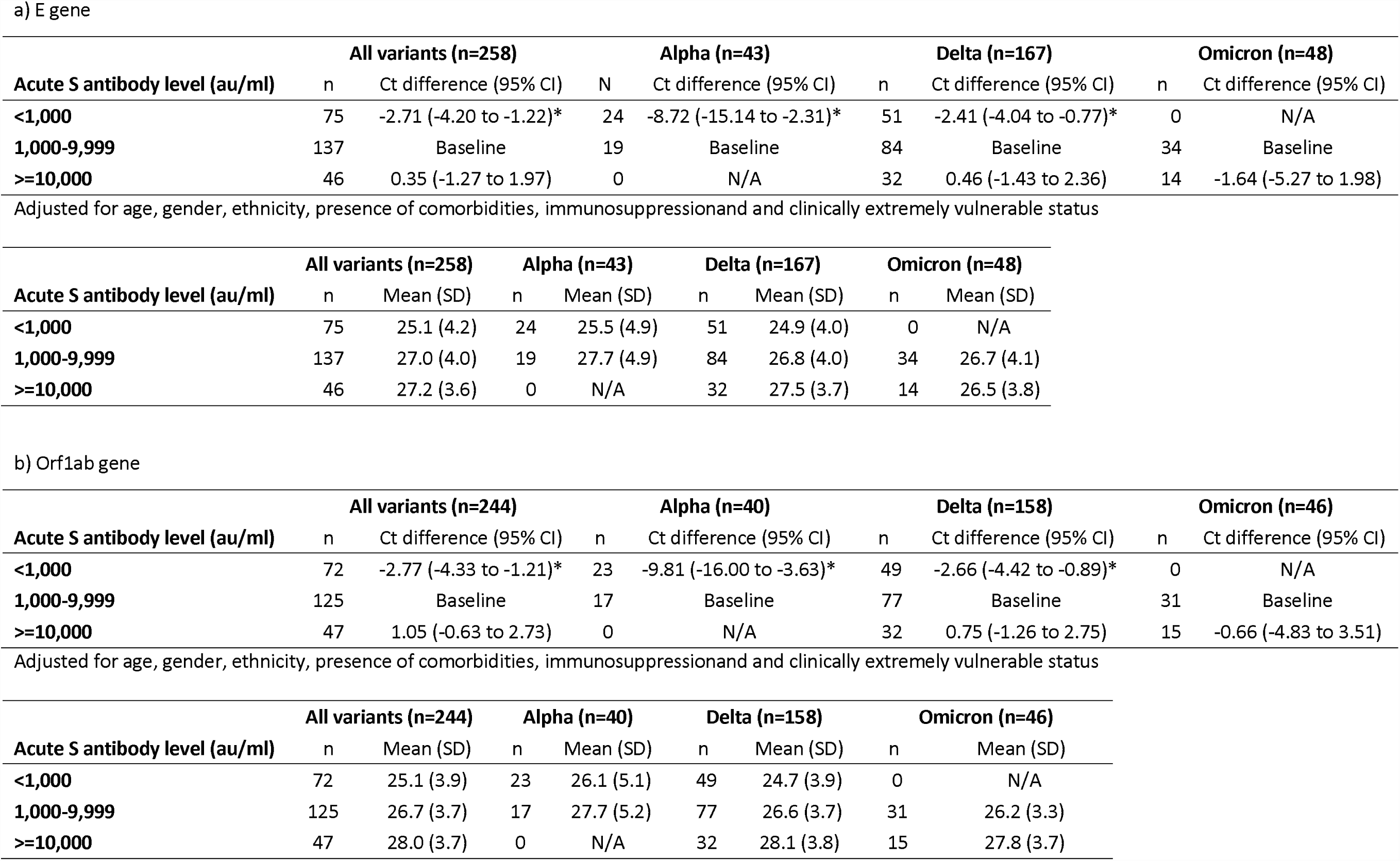
Random effects linear regression assessing the effect of anti-S antibody level in acute sera on Ct values and mean Ct values from both a)E gene and b)Orf1ab gene regions over the course of a SARS-Cov-2 infection by all variants combined and by variants in vaccinated individuals.

Overall and when stratified by variant, there was no significant association between the acute anti-S antibody level and self-reported symptom severity (graded as mild versus moderate or severe; odds ratio (OR)<1: less likely to be severe) (Table 7). The OR of having moderate/severe disease was lower in individuals with a level of 10,000 au/ml or more acute anti-S antibody compared to individuals with 1,000 to 9,999 au/ml anti-S antibody level but the differences were not significant. Overall and for Delta, the OR of reporting six or more symptoms was significantly lower in individuals with a level of 10,000 au/ml or more acute anti-S antibody compared to individuals with 1,000 to 9,999 au/ml anti-S antibody level (OR (95% CI): Overall: 0.15 (0.04-0.49), Delta: 0.12 (0.03-0.55)), for Omicron, the OR was also lower but not significant. There was no significant difference for the Alpha variant. (Table 8).

**Table 7.**
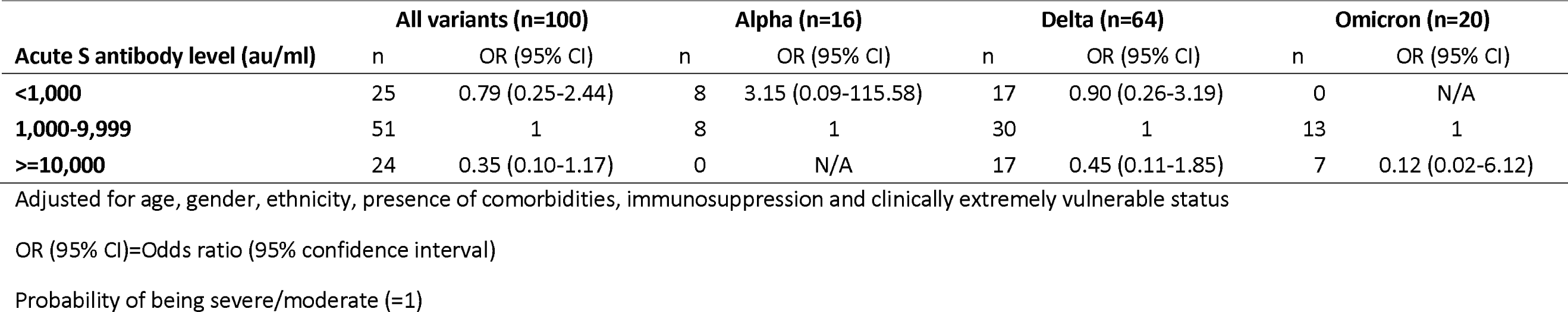
Multivariable logistic regression assessing the effect of anti-S antibody level in acute sera on self-reported symptom severity (mild vs moderate/severe) of SARS-Cov-2 infection by all variants combined and by variants in vaccinated individuals.

| **Acute S antibody level (au/ml)** | **All variants (n=100)** |  | **Alpha (n=16)** |  | **Delta (n=64)** |  | **Omicron (n=20)** |  |
| --- | --- | --- | --- | --- | --- | --- | --- | --- |
|  | n | OR (95% CI) | n | OR (95% CI) | n | OR (95% CI) | n | OR (95% CI) |
| **<1,000** | 25 | 0.79 (0.25-2.44) | 8 | 3.15 (0.09-115.58) | 17 | 0.90 (0.26-3.19) | 0 | N/A |
| **1,000-9,999** | 51 | 1 | 8 | 1 | 30 | 1 | 13 | 1 |
| **>=10,000** | 24 | 0.35 (0.10-1.17) | 0 | N/A | 17 | 0.45 (0.11-1.85) | 7 | 0.12 (0.02-6.12) |

Adjusted for age, gender, ethnicity, presence of comorbidities, immunosuppression and clinically extremely vulnerable status
OR (95% CI)=Odds ratio (95% confidence interval)
Probability of being severe/moderate (=1)

**Table 8.**
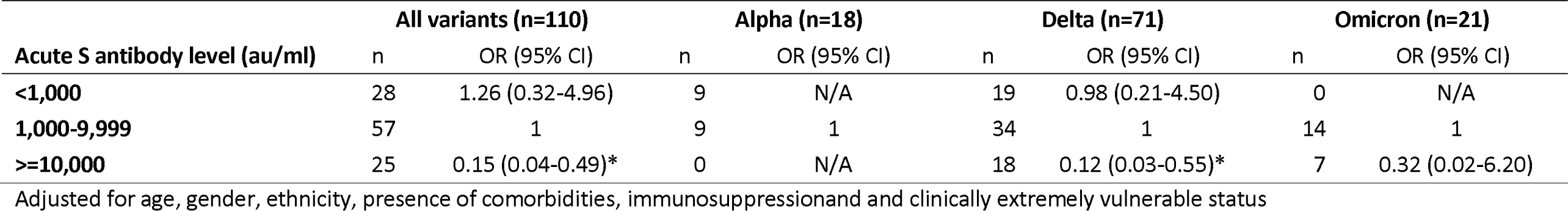
Multivariable logistic regression assessing the effect of anti-S antibody level in acute sera on self-reported number of symptoms (1-5 vs 6≥ symptoms) of SARS-Cov-2 infection, by all variants and variants in vaccinated individuals.

## Discussion

### Summary of results

Following infection, convalescent anti-N antibody levels were lower in vaccinated individuals and convalescent anti-S antibody levels were higher in vaccinated individuals and increased with the number of doses received. Moreover, as expected, the acute anti-S antibody level increased with the number of doses received. Shorter duration of positivity and higher mean Ct values (indicating a lower viral load) were observed with increasing acute anti-S antibody for all variants, this was statistically significant overall and for Delta and for Alpha for the mean Ct value. Individuals with higher acute anti-S antibody level were less likely to report moderate or severe symptoms for all variants. Individuals with higher acute anti-S antibody level were less likely to report six or more symptoms overall and for all variants (statistically significant for Delta). Generally, a lower antibody response was observed for immunosuppressed and CEV individuals.

### Interpretation and comparison with other studies

The higher convalescent anti-S antibody levels in vaccinated individuals is consistent with other studies (20) whereas few studies shown a lower convalescent anti-N antibody levels in vaccinated individuals (21).

The lower convalescent anti-N and acute anti-S antibody response (not significant for acute anti-S) in immunosuppressed individuals is concordant with other studies (22, 23) and is likely due to specific health conditions or medical treatments which may weaken the immune system and impair the ability to generate a strong immune response to COVID-19 vaccination or infection.

Overall and for the Delta variant, the convalescent anti-N antibody level was higher in other ethnic groups than in the white ethnic group. Higher antibody levels in individuals of other ethnic groups have been reported in other studies (24, 25). In accordance with other studies, we also found that the convalescent anti-N antibody level was higher in males (7, 24, 25). Older age has been reported to be associated to higher antibody levels (7, 24, 25) but this was less obvious in our study.

We found that higher acute anti-S antibody level was associated with a reduced duration of positivity and lower viral load. This finding suggests a lower risk of onwards transmission which is consistent with previous transmission studies that shown similar associations (26). Antibody levels have been correlated with protection (27, 28) and this finding suggests that those with higher antibody levels may have milder disease as they clear the infection more rapidly. We did not find a significant association between the acute anti-S antibody levels and self-reported symptom severity, but we did find an association with the number of symptoms reported - individuals with higher acute anti-S antibody levels reported fewer symptoms overall, and for the Delta variant (although this was not observed for Alpha and Omicron). It is possible that no association with symptom severity was observed as this is a highly subjective measure relying on an individuals’ own perception of their illness. The ratio of baseline to convalescent anti-N antibody levels following infection was greater for unvaccinated vs vaccinated individuals; this may also reflect a milder infection occurring in vaccinated individuals which did not provoke as strong anti-N antibody response.

Significant differences were sometimes observed for the Delta variant, but not for Alpha or Omicron. Our study started in February 2021 and ended in April 2022, covering a long period where Delta was dominant, most individuals recruited were infected with Delta. With a greater sample size, we were better powered to detect smaller differences for Delta than we were for either Alpha or Omicron.

Other parts of the immune system, not measured in this study, also contribute to the response to SARS-CoV-2 infection. Memory B cells and T cells are immune cells that play a role in protecting against SARS-CoV-2 infection as they may be maintained and provide long term protection against the infection as antibody response wane (29, 30).

### Strengths and limitations

This is one of the most detailed studies to investigate antibody response by variant in vaccine eligible individuals alongside detailed virological and clinical data. This level of detail alongside the continuous nature of the study over a period during which three major variants emerged in the UK is a significant strength. Nevertheless, there are also several limitations. Despite successfully recruiting a large number of participants, the recruitment was based on testing positive and may not be representative of the whole population as people tested might be different to people who don’t want to be tested. Moreover, despite having a large number of participants, we could not look at intervals post vaccination or manufacturer data in details. In addition, self-reported information is prone to recall bias and can lead to misclassification as there may be a difference in the way individuals remember or report information. This misclassification would have to vary according to vaccination status or variant to affect the results. During the enhanced surveillance study there were many changes in testing policies, testing eligibility, vaccine offer eligibility, social-distancing behaviours and national lockdown policies which affected who was recruited into the study between the different variant periods. Unvaccinated individuals recruited during the Omicron period could have different characteristics of those recruited during the Alpha and Delta periods, since some were not eligible for vaccination yet during the Alpha and Delta periods but by Omicron all adults would have been eligible. Freely available PCR testing in the community ended at the end of March 2022, so any individuals recruited after this period may have different characteristics to those recruited before. In addition, recruits who received three doses of vaccines are more likely to have different attributes than recruits who received less than three doses of vaccines (e.g., older, immunosuppressed, CEV, etc.) due to higher risk individuals being prioritised for booster vaccination during the recruitment period of the study. To take these attributes into account, we used statistical methods that allowed us to account for multiple potential confounding factors, though residual confounding may still affect our findings.

In the analysis sample, we excluded individuals with a prior infection based on a prior positive anti-N antibody result or self-reported previous positive PCR, or antibody test at recruitment. It’s possible that we did not exclude all the individuals with a prior infection if these individuals had not developed antibodies at the time of the test or if they had too low level of antibodies to reach the level that the test calls positive.

### Conclusion

This study furthers our understanding of the anti-N and anti-S antibody response following infection by vaccination status. We find that vaccination increases convalescent anti-S antibody level but reduces convalescent anti-N antibody level. Furthermore, we were able to find evidence of an association between high acute anti-S antibody level and reduced duration of positivity and increased Ct values (lower viral load), suggesting a lower risk of onwards transmission. High anti-S antibody level may also be associated with reduced number of symptoms and symptoms severity. Our finding supports other real-world vaccine effectiveness studies demonstrating the benefits of vaccination on symptom severity following infection.

## Data Availability

Data cannot be made publicly available for ethical and legal reasons, i.e. public availability would compromise patient confidentiality as data tables list single counts of individuals rather than aggregated data

## Acknowledgements

We thank all participants for enrolling within the Enhanced Surveillance study and providing samples. We also thank all staff that enable the Enhanced surveillance to operate and Antoaneta Bukasa, Charlotte Gower. We thank the UKHSA team of research nurses: Louise Letley, Donna Haskins, Catriona Angel, Skye Firminger, Kay Ratcliffe, Angela Sherridan, Shelina Rajan, Lola Akindele. We thank those who managed and assembled the kits for the study: Deborah Cohen, Teresa Gibbs, Kim Taylor. We thank the UKHSA reference laboratory team who tested all samples: Samreen Ijaz, Maria Zambon, Kevin Brown. We thank the Molis team who enabled us to access the study results.

## Author’s contributions

CQ, RL, FK, CS, LL, DH, CA, SF, KR, AS, SR, LA were responsible for aspects of recruitment and data collection. CQ, RL, FK, CS were responsible for data management. JLB conceived the study. JLB, FK and CG created the study design. CQ designed the analysis plan and performed statistical analysis with input from NA, HW, FK, JLB. CQ and RL produced the data cleaning script for the underlying dataset. CQ, RL and FK did the data linkage. CQ and RL accessed and verified the underlying data. CQ wrote the first draft of the manuscript. All authors contributed to and reviewed the final submitted manuscript.

## Ethics

Enhanced surveillance protocols have been approved by the UKHSA Research Ethics and Governance of Public Health Practice Group (REGG). Informed consent was obtained from participants. All study participants provided oral informed consent for sampling and follow up and informed they were free to decline to participate in the surveillance at any time.

## References

1. Agency UHS. Guidance COVID-19: the green book, chapter 14a 2023 [Available from: https://www.gov.uk/government/publications/covid-19-the-green-book-chapter-14a.

2. GOV.UK. Investigation of SARS-CoV-2 variants: technical briefings [Available from: https://www.gov.uk/government/publications/investigation-of-sars-cov-2-variants-technical-briefings.

3. Twohig KA, Nyberg T, Zaidi A, Thelwall S, Sinnathamby MA, Aliabadi S, et al. Hospital admission and emergency care attendance risk for SARS-CoV-2 delta (B.1.617.2) compared with alpha (B.1.1.7) variants of concern: a cohort study. Lancet Infect Dis. 2022;22(1):35–42.

4. Nyberg T, Ferguson NM, Nash SG, Webster HH, Flaxman S, Andrews N, et al. Comparative analysis of the risks of hospitalisation and death associated with SARS-CoV-2 omicron (B.1.1.529) and delta (B.1.617.2) variants in England: a cohort study. Lancet (London, England). 2022;399(10332):1303–12.

5. GOV.UK. UK COVID-19 vaccines delivery plan [Available from: https://www.gov.uk/government/publications/uk-covid-19-vaccines-delivery-plan/uk-covid-19-vaccines-delivery-plan.

6. Jiang S, Hillyer C, Du L. Neutralizing Antibodies against SARS-CoV-2 and Other Human Coronaviruses: (Trends in Immunology 41, 355-359; 2020). Trends in immunology. 2020;41(6):545.

7. Navaratnam AMD, Shrotri M, Nguyen V, Braithwaite I, Beale S, Byrne TE, et al. Nucleocapsid and spike antibody responses following virologically confirmed SARS-CoV-2 infection: an observational analysis in the Virus Watch community cohort. Int J Infect Dis. 2022;123:104–11.

8. Krutikov M, Palmer T, Tut G, Fuller C, Azmi B, Giddings R, et al. Prevalence and duration of detectable SARS-CoV-2 nucleocapsid antibodies in staff and residents of long-term care facilities over the first year of the pandemic (VIVALDI study): prospective cohort study in England. Lancet Healthy Longev. 2022;3(1):e13–e21.

9. Tut G, Lancaster T, Sylla P, Butler MS, Kaur N, Spalkova E, et al. Antibody and cellular immune responses following dual COVID-19 vaccination within infection-naive residents of long-term care facilities: an observational cohort study. Lancet Healthy Longev. 2022;3(7):e461–e9.

10. Shrotri M, Harris RJ, Rodger A, Planche T, Sanderson F, Mahungu T, et al. Persistence of SARS-CoV-2 N-Antibody Response in Healthcare Workers, London, UK. Emerging infectious diseases. 2021;27(4):1155–8.

11. Vusirikala A, Whitaker H, Jones S, Tessier E, Borrow R, Linley E, et al. Seroprevalence of SARS-CoV-2 antibodies in university students: Cross-sectional study, December 2020, England. The Journal of infection. 2021;83(1):104–11.

12. Siggins MK, Thwaites RS, Openshaw PJM. Durability of Immunity to SARS-CoV-2 and Other Respiratory Viruses. Trends in microbiology. 2021;29(7):648–62.

13. Gobbi F, Buonfrate D, Moro L, Rodari P, Piubelli C, Caldrer S, et al. Antibody Response to the BNT162b2 mRNA COVID-19 Vaccine in Subjects with Prior SARS-CoV-2 Infection. Viruses. 2021;13(3).

14. Aldridge RW, Yavlinsky A, Nguyen V, Eyre MT, Shrotri M, Navaratnam AMD, et al. SARS-CoV-2 antibodies and breakthrough infections in the Virus Watch cohort. Nat Commun. 2022;13(1):4869.

15. GOV.UK. SIREN study [Available from: https://www.gov.uk/guidance/siren-study.

16. Tessier E, Edelstein M, Tsang C, Kirsebom F, Gower C, Campbell CNJ, et al. Monitoring the COVID-19 immunisation programme through a national immunisation Management system - England’s experience. International journal of medical informatics. 2023;170:104974.

17. Hoschler K, Ijaz S, Andrews N, Ho S, Dicks S, Jegatheesan K, et al. SARS Antibody Testing in Children: Development of Oral Fluid Assays for IgG Measurements. Microbiology spectrum. 2022;10(1):e0078621.

18. Ijaz S, Dicks S, Jegatheesan K, Parker E, Katsanovskaja K, Vink E, et al. Mapping of SARS-CoV-2 IgM and IgG in gingival crevicular fluid: Antibody dynamics and linkage to severity of COVID-19 in hospital inpatients. The Journal of infection. 2022;85(2):152–60.

19. Singanayagam A, Patel M, Charlett A, Lopez Bernal J, Saliba V, Ellis J, et al. Duration of infectiousness and correlation with RT-PCR cycle threshold values in cases of COVID-19, England, January to May 2020. 2020;25(32):2001483.

20. Farnsworth CW, O’Neil CA, Dalton C, McDonald D, Vogt L, Hock K, et al. Association between SARS-CoV-2 Symptoms, Ct Values, and Serological Response in Vaccinated and Unvaccinated Healthcare Personnel. The journal of applied laboratory medicine. 2023.

21. Whitaker HJ, Gower C, Otter AD, Simmons R, Kirsebom F, Letley L, et al. Nucleocapsid antibody positivity as a marker of past SARS-CoV-2 infection in population serosurveillance studies: impact of variant, vaccination, and choice of assay cut-off. 2021:2021.10.25.21264964.

22. Whitaker HJ, Tsang RSM, Byford R, Andrews NJ, Sherlock J, Sebastian Pillai P, et al. Pfizer-BioNTech and Oxford AstraZeneca COVID-19 vaccine effectiveness and immune response amongst individuals in clinical risk groups. The Journal of infection. 2022;84(5):675–83.

23. Shrotri M, Fragaszy E, Nguyen V, Navaratnam AMD, Geismar C, Beale S, et al. Spike-antibody responses to COVID-19 vaccination by demographic and clinical factors in a prospective community cohort study. Nat Commun. 2022;13(1):5780.

24. Lumley SF, Wei J, O’Donnell D, Stoesser NE, Matthews PC, Howarth A, et al. The Duration, Dynamics, and Determinants of Severe Acute Respiratory Syndrome Coronavirus 2 (SARS-CoV-2) Antibody Responses in Individual Healthcare Workers. Clinical infectious diseases : an official publication of the Infectious Diseases Society of America. 2021;73(3):e699–e709.

25. Post N, Eddy D, Huntley C, van Schalkwyk MCI, Shrotri M, Leeman D, et al. Antibody response to SARS-CoV-2 infection in humans: A systematic review. PLoS One. 2020;15(12):e0244126.

26. Clifford S, Waight P, Hackman J, Hué S, Gower CM, Kirsebom FC, et al. Effectiveness of BNT162b2 and ChAdOx1 against SARS-CoV-2 household transmission: a prospective cohort study in England. 2021:2021.11.24.21266401.

27. Wei J, Pouwels KB, Stoesser N, Matthews PC, Diamond I, Studley R, et al. Antibody responses and correlates of protection in the general population after two doses of the ChAdOx1 or BNT162b2 vaccines. Nat Med. 2022;28(5):1072–82.

28. Wei J, Matthews PC, Stoesser N, Newton JN, Diamond I, Studley R, et al. Protection against SARS-CoV-2 Omicron BA.4/5 variant following booster vaccination or breakthrough infection in the UK. Nature Communications. 2023;14(1):2799.

29. Moss P. The T cell immune response against SARS-CoV-2. Nature immunology. 2022;23(2):186–93.

30. Gaebler C, Wang Z, Lorenzi JCC, Muecksch F, Finkin S, Tokuyama M, et al. Evolution of antibody immunity to SARS-CoV-2. Nature. 2021;591(7851):639–44.

