## supplementary materials for "Serological outcomes of SARS-CoV-2 infection by vaccination status and variant in England"

Supplementary Figures and Tables (pages 2-15)

Supplementary materials for the methods section – description of the data sources (pages 16-17)

**Supplementary Figures and Tables**

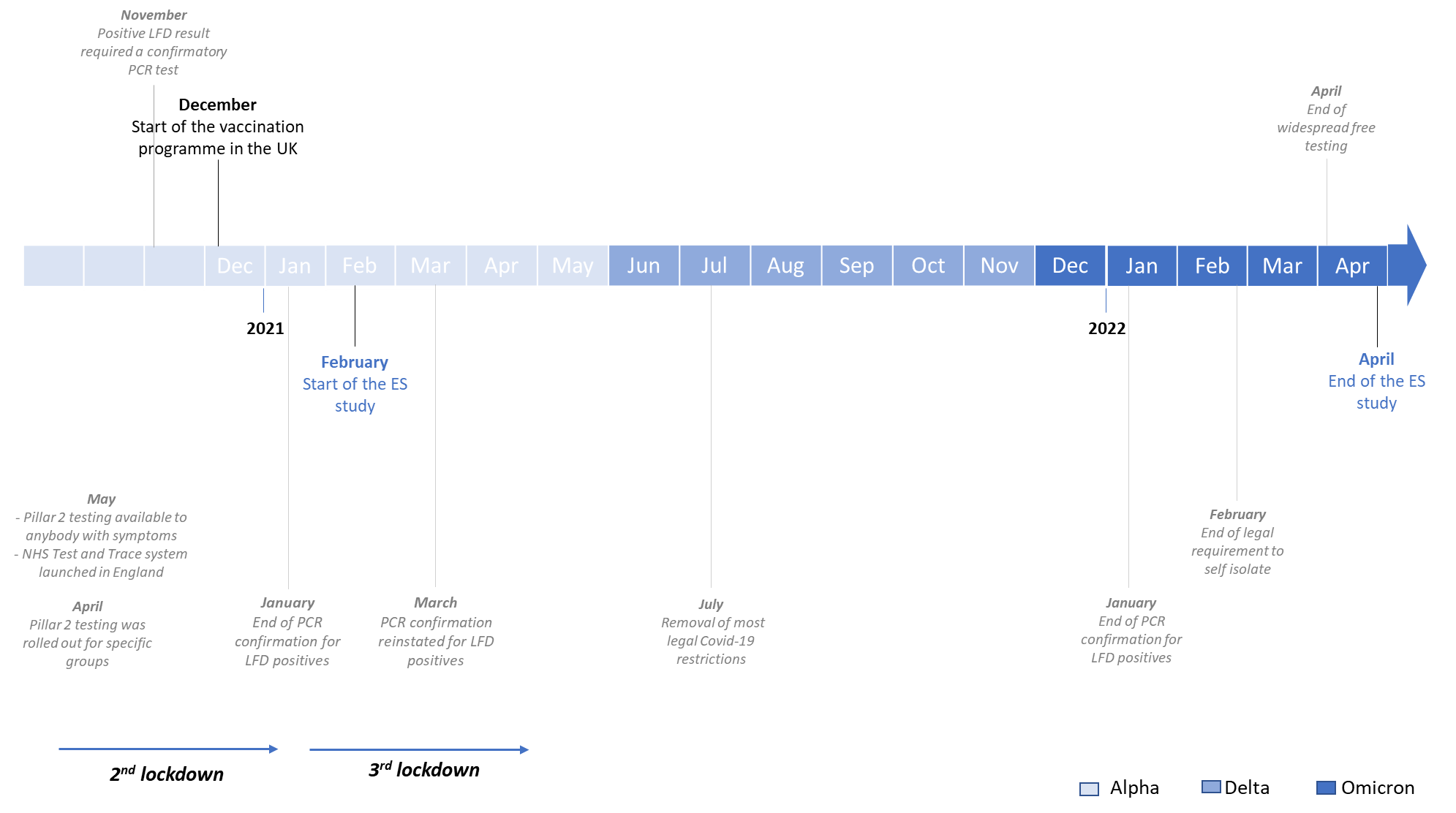

Figure 1. COVID-19 timeline in United Kingdom from 2020 to 2022

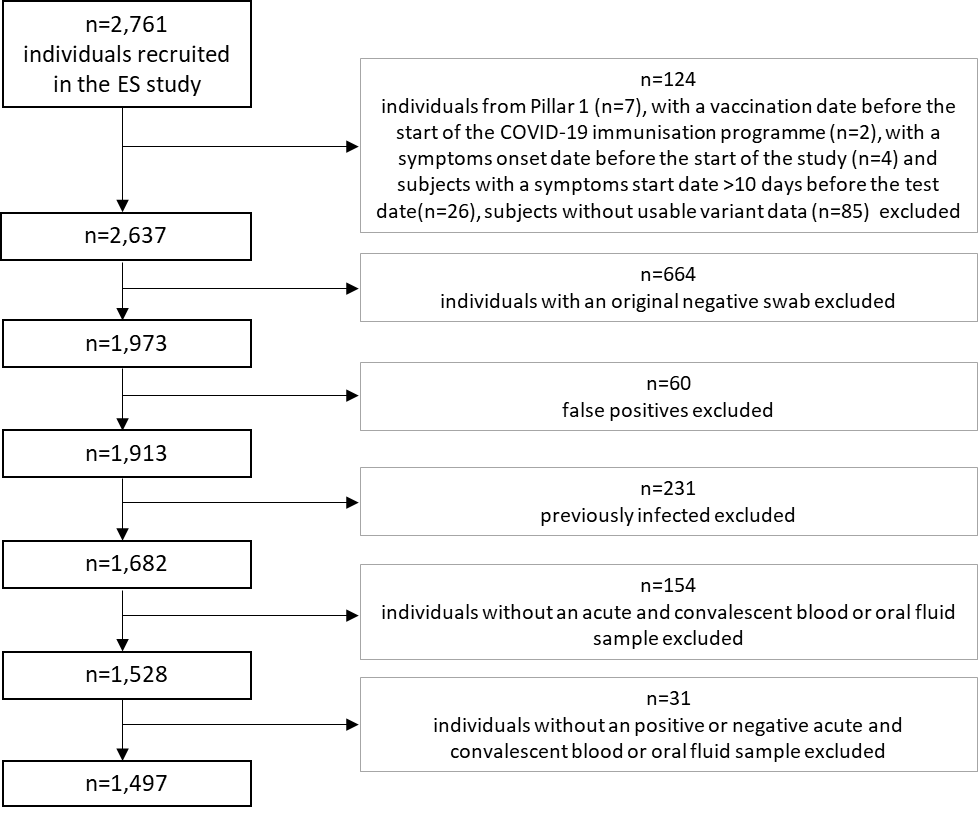

Figure 2. Flow chart of the individuals from the enhanced surveillance (ES) study of COVID-19 vaccines in England included in the analysis

Figure 3. Enhanced surveillance questionnaire at recruitment (1/2)

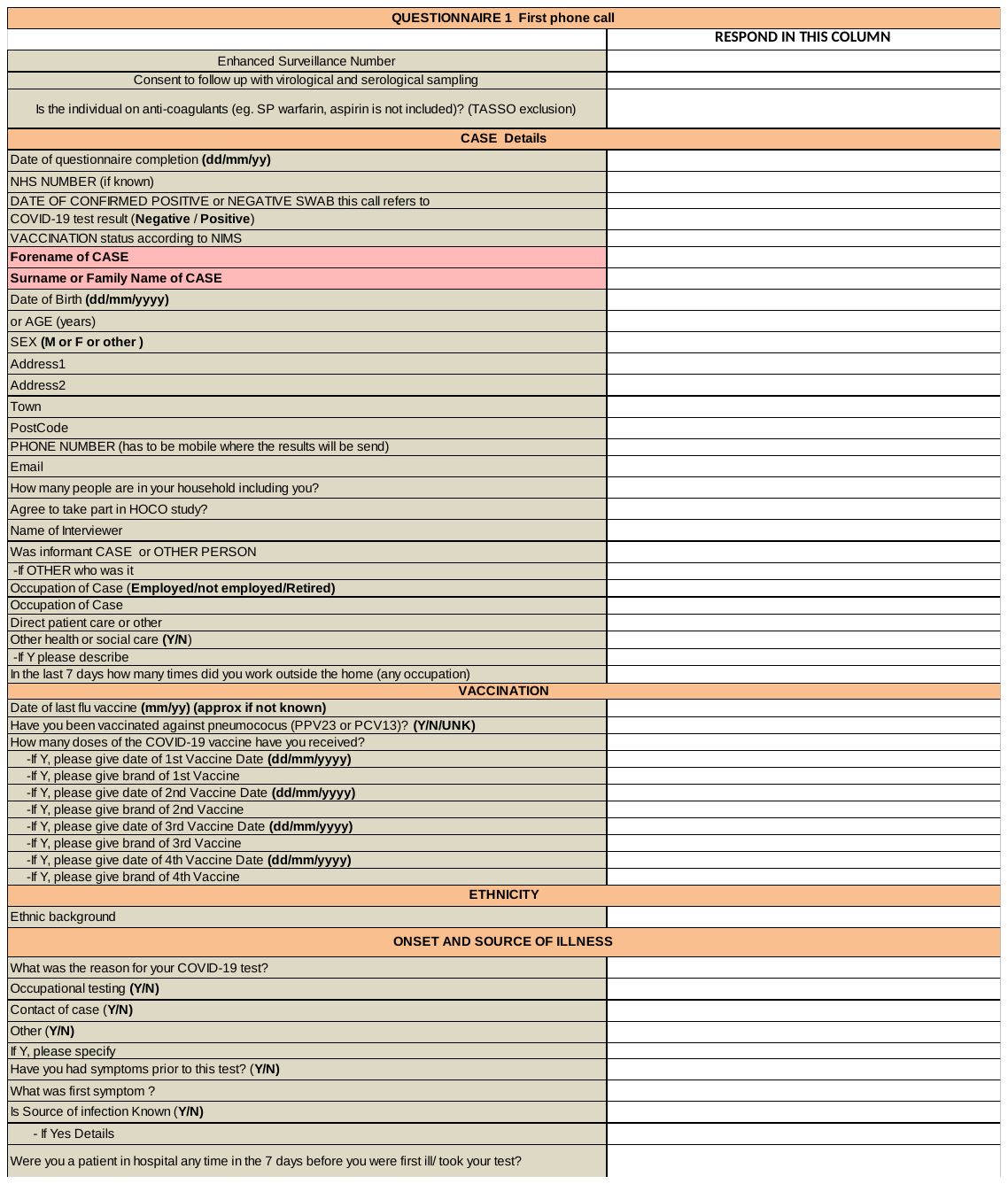

Enhanced surveillance questionnaire at recruitment (2/2)

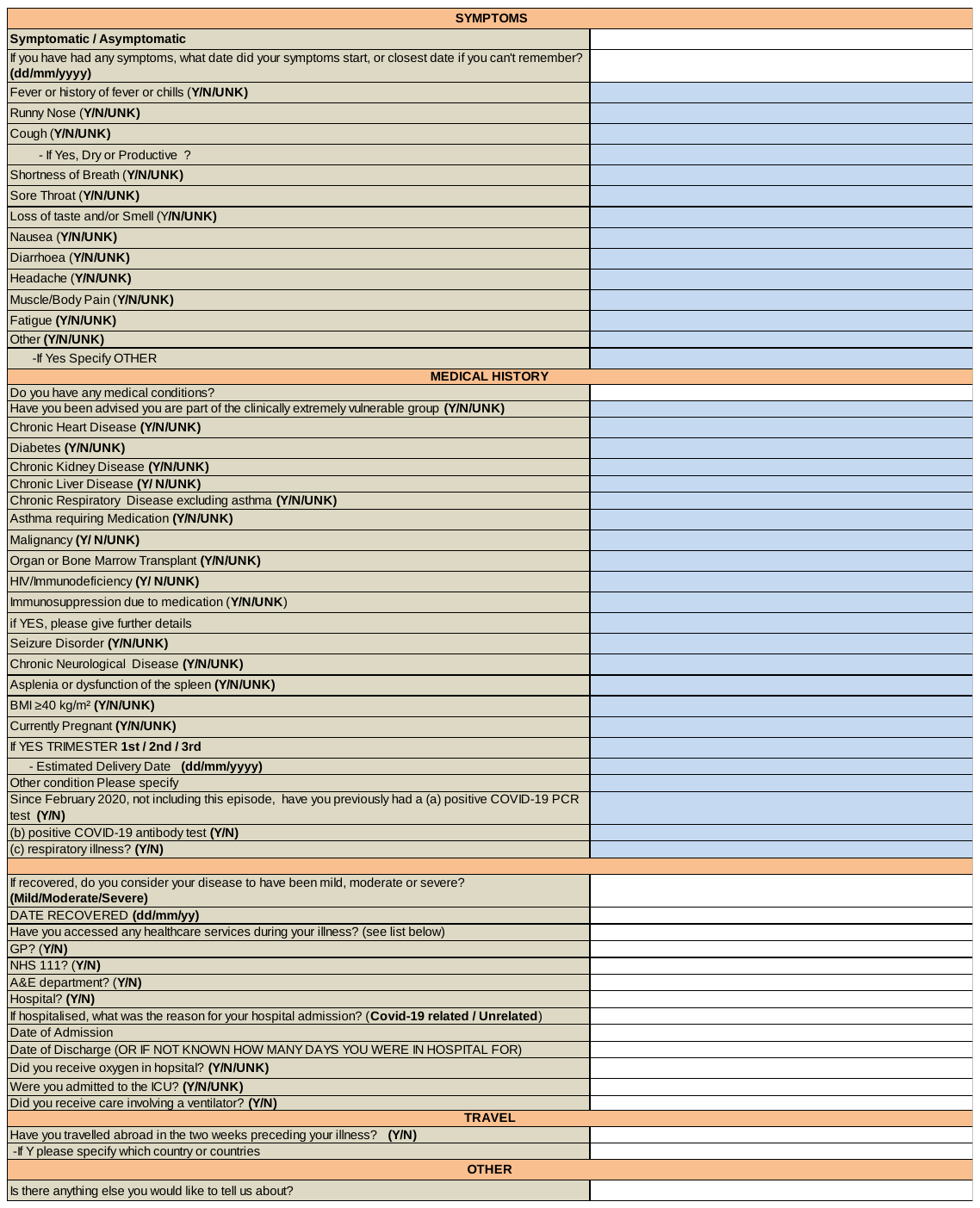

Enhanced surveillance questionnaire 21 days post recruitment

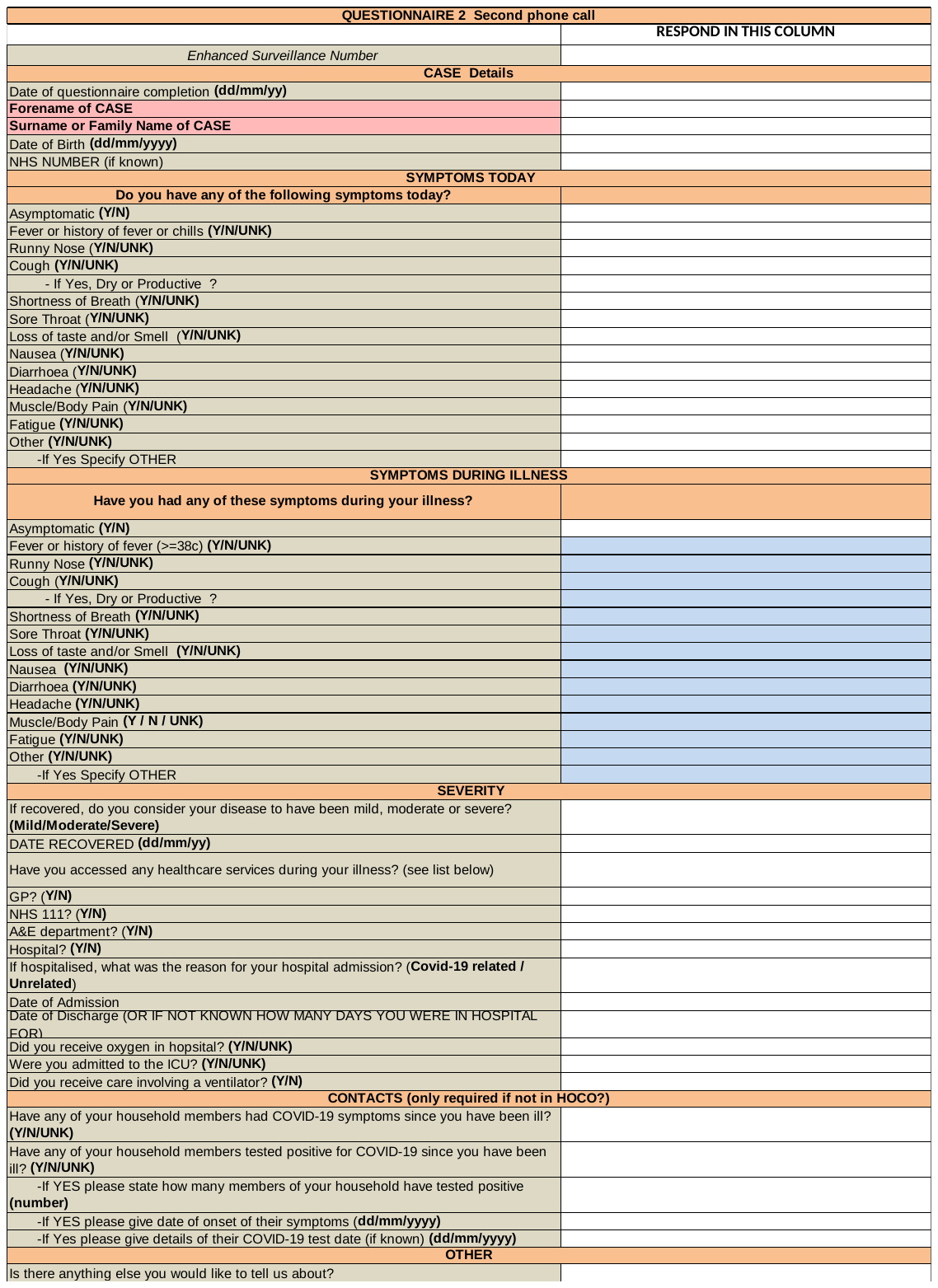

Table 1.Description of the number of self-reported symptoms and self-reported specific symptoms by vaccination status and variants, N=1,464

|  | **Alpha** | | | **Delta** | | | | **Omicron** | | | **Overall** |
| --- | --- | --- | --- | --- | --- | --- | --- | --- | --- | --- | --- |
|  | Unvaccinated | >28 days post D1 | >14 days post D2 | Unvaccinated | >28 days post D1 | >14 days post D2 | >14 days post D3+ | Unvaccinated | >14 days post D2 | >14 days post D3+ |  |
|  | n (%) | n (%) | n (%) | n (%) | n (%) | n (%) | n (%) | n (%) | n (%) | n (%) |  |
| **Number of symptoms** |  |  |  |  |  |  |  |  |  |  |  |
| 1-3 | 13 (6.2) | 11 (8.9) | 1 (6.3) | 8 (6.7) | 10 (18.9) | 33 (6.5) | 15 (22.1) | 5 (14.3) | 4 (6.0) | 18 (6.8) | 118 (8.1) |
| 4-5 | 27 (12.9) | 26 (21.0) | 5 (31.2) | 11 (9.2) | 3 (5.7) | 80 (15.7) | 10 (14.7) | 3 (8.6) | 7 (10.5) | 46 (17.4) | 218 (14.9) |
| 6-7 | 58 (27.8) | 37 (29.8) | 3 (18.8) | 21 (17.7) | 12 (22.6) | 116 (22.8) | 8 (11.8) | 19 (25.7) | 22 (32.8) | 85 (32.2) | 371 (25.3) |
| ≥8 | 111 (53.1) | 50 (40.3) | 7 (43.7) | 79 (66.4) | 28 (52.8) | 280 (55.0) | 35 (51.4) | 18 (51.4) | 34 (50.7) | 115 (43.6) | 757 (51.7) |
| **Specific symptoms** |  |  |  |  |  |  |  |  |  |  |  |
| Fever | 129 (61.7) | 57 (46.0) | 8 (50.0) | 84 (70.6) | 32 (60.4) | 316 (62.1) | 25 (36.8) | 29 (82.9) | 45 (67.2) | 136 (51.5) | 861 (58.8) |
| Runny nose | 118 (56.5) | 87 (70.2) | 10 (62.5) | 60 (50.4) | 39 (73.6) | 387 (76.0) | 58 (85.3) | 19 (54.3) | 48 (71.6) | 206 (78.0) | 1,032 (70.5) |
| Cough | 178 (85.2) | 101 (81.5) | 8 (50.0) | 104 (87.4) | 43 (81.1) | 415 (81.5) | 41 (60.3) | 27 (77.1) | 56 (83.6) | 232 (87.9) | 1,205 (82.3) |
| Shortness of breath | 89 (42.6) | 54 (43.6) | 7 (43.8) | 50 (42.0) | 20 (37.7) | 182 (35.8) | 29 (42.7) | 11 (31.4) | 22 (32.8) | 101 (38.3) | 565 (38.6) |
| Sore throat | 116 (55.5) | 62 (50.0) | 4 (25.0) | 65 (54.6) | 23 (43.4) | 283 (55.6) | 27 (39.7) | 17 (48.6) | 48 (71.6) | 183 (69.3) | 828 (56.5) |
| Loss of taste and smell | 125 (59.8) | 65 (52.4) | 12 (75.0) | 88 (73.9) | 38 (71.7) | 372 (73.1) | 38 (55.9) | 15 (42.9) | 19 (28.4) | 76 (28.8) | 848 (57.9) |
| Nausea | 73 (34.9) | 25 (20.2) | 4 (25.0) | 54 (45.4) | 20 (37.7) | 126 (24.7) | 18 (26.5) | 11 (31.4) | 24 (35.8) | 54 (20.5) | 409 (27.9) |
| Diarrhoea | 55 (26.3) | 24 (19.4) | 3 (18.8) | 33 (27.5) | 11 (20.8) | 101 (19.8) | 13 (19.1) | 14 (40.0) | 16 (23.9) | 51 (19.3) | 321 (21.9) |
| Headache | 168 (80.4) | 97 (78.2) | 14 (87.5) | 100 (84.0) | 42 (79.2) | 402 (79.0) | 52 (76.5) | 26 (74.2) | 58 (86.6) | 198 (75.0) | 1,157 (79.0) |
| Pain | 164 (78.5) | 77 (62.1) | 9 (56.3) | 94 (79.0) | 32 (60.4) | 344 (67.6) | 34 (50.0) | 23 (65.7) | 51 (76.1) | 159 (60.2) | 987 (67.4) |
| Fatigue | 181 (86.6) | 99 (79.8) | 15 (93.8) | 100 (84.0) | 48 (90.6) | 427 (83.9) | 51 (75.0) | 25 (71.4) | 60 (89.6) | 225 (85.2) | 1,231 (84.1) |
| Other | 139 (66.5) | 78 (62.9) | 13 (81.3) | 87 (73.1) | 33 (62.3) | 356 (69.9) | 49 (72.1) | 26 (74.3) | 41 (61.2) | 181 (68.6) | 1,003 (68.5) |
| Total | 209 (100.0) | 124 (100.0) | 16 (100.0) | 119 (100.0) | 53 (100.0) | 509 (100.0) | 68 (100.0) | 35 (100.0) | 67 (100.0) | 264 (100.0) | 1,464 (100.0) |

Table 2.Description of the number of acute and convalescent blood and oral fluid samples included in the analysis for Roche N and Roche S (qualitative results)

|  |  | **Serum sample** | | **Oral fluid sample** | | **Serum and Oral fluid samples** | |
| --- | --- | --- | --- | --- | --- | --- | --- |
|  |  | **Acute**  **n (%)** | **Convalescent**  **n (%)** | **Acute**  **n (%)** | **Convalescent**  **n (%)** | **Acute**  **n (%)** | **Convalescent**  **n (%)** |
| **Anti-N antibody level** | Positive | 0 (0.0) | 1,201 (90.6) | 0 (0.0) | 88 (57.5) | 0 (0.0) | 1,236 (92.7) |
|  | Negative | 671 (47.7) | 42 (3.2) | 730 (87.4) | 32 (20.9) | 1,308 (89.6) | 58 (4.4) |
|  | Equivocal | 25 (1.8) | 23 (1.7) | 6 (0.7) | 8 (5.2) | 31 (2.1) | 23 (1.7) |
|  | Insufficient | 708 (50.3) | 57 (4.3) | 76 (9.1) | 23 (15.0) | 119 (8.2) | 13 (1.0) |
|  | Not tested | 4 (0.3) | 2 (0.2) | 24 (2.9) | 2 (1.3) | 2 (0.1) | 3 (0.2) |
|  | Total | 1,408 (100.0) | 1,325 (100.0) | 836 (100.0) | 153 (100.0) | 1,460 (100.0) | 1,333 (100.0) |
| **Anti-S antibody level** | Positive | 357 (37.2) | 1,259 (95.0) | 466 (55.8) | 119 (77.8) | 783 (66.0) | 1,307 (98.0) |
|  | Negative | 73 (7.3) | 7 (0.5) | 261 (31.3) | 9 (5.9) | 289 (24.3) | 10 (0.8) |
|  | Equivocal | 0 (0.0) | 0 (0.0) | 8 (1.0) | 0 (0.0) | 8 (0.7) | 0 (0.0) |
|  | Insufficient | 527 (55.0) | 57 (4.3) | 76 (9.1) | 23 (15.0) | 107 (9.0) | 13 (1.0) |
|  | Not tested | 2 (0.2) | 2 (0.2) | 24 (2.9) | 2 (1.3) | 0 (0.0) | 3 (0.2) |
|  | Total | 959 (100.0) | 1,325 (100.0) | 835 (100.0) | 153 (100.0) | 1,187 (100.0) | 1,333 (100.0) |

Positive acute N samples excluded

A blood sample was defined as acute for anti-N antibody if the interval between the sample date and the symptom onset date or the date of original PCR positive test was less than 10 days. A blood sample was defined as acute for anti-S antibody if the interval between the sample date and the test or symptom date was less than 6 days. A blood sample was defined to be convalescent for anti-N and anti-S antibodies if the interval between the sample date and the test or symptom date was more than 21 days.

Table 3. Description of the acute and convalescent blood sample with quantitative results included in the analysis

|  | **Anti-N antibody level** | | | | | | **Anti-S antibody level** | | | | | |
| --- | --- | --- | --- | --- | --- | --- | --- | --- | --- | --- | --- | --- |
|  | n | Quartile 1 | Median | Quartile 3 | Minimum | Maximum | n | Quartile 1 | Median | Quartile 3 | Minimum | Maximum |
| **Acute** | 696 | 0.07 | 0.08 | 0.10 | 0.05 | 0.97 | 430 | 248 | 2,966.5 | 8,706 | 0.4 | 128,900 |
| **Convalescent** | 1,266 | 5.60 | 16.60 | 45.20 | 0.07 | 267 | 1,266 | 3,507 | 16,474.5 | 46,900 | 0.4 | 342,900 |

Based on results from serum samples only. It was not possible to combine with the oral fluid quantitative results as it’s not the same information.

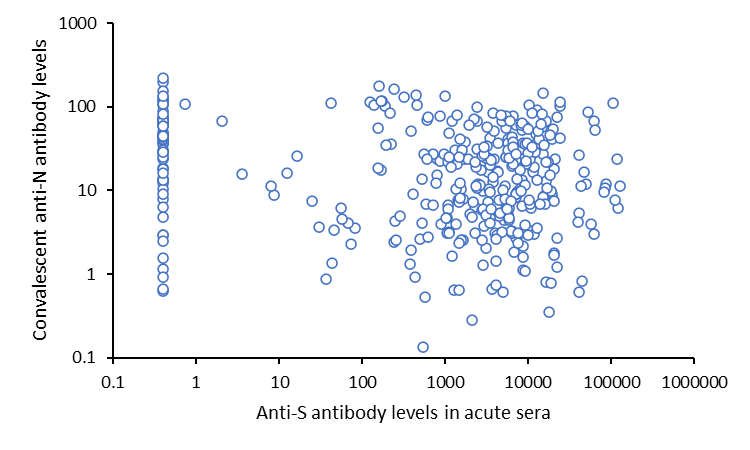

Figure 4. Convalescent anti-N antibody levels according to acute anti-S antibody levels

Table 4. Description of the convalescent anti-N antibody seroconversion proportion by vaccination status and variants

|  |  | | **Alpha** | | | | | | | **Delta** | | | | | | | | | **Omicron** | | | | | | |
| --- | --- | --- | --- | --- | --- | --- | --- | --- | --- | --- | --- | --- | --- | --- | --- | --- | --- | --- | --- | --- | --- | --- | --- | --- | --- |
|  |  |  | **Unvaccinated** | | **>28 days post D1** | | **>14 days post D2** | | **TOTAL** | **Unvaccinated** | | **>28 days post D1** | | **>14 days post D2** | | **>14 days post D3+** | | **TOTAL** | **Unvaccinated** | | **>14 days post D2** | | **>14 days post D3+** | | **TOTAL** |
|  |  |  | n | % | n | % | n | % | **n** | n | % | n | % | n | % | n | % | **n** | n | % | n | % | n | % | **n** |
| **Seroconversion** | | Yes | 155 | 94.5 | 101 | 93.5 | 16 | 94.1 | **272** | 82 | 95.3 | 42 | 97.7 | 384 | 97.2 | 49 | 96.1 | **557** | 23 | 92.0 | 42 | 95.5 | 171 | 94.0 | **236** |
|  |  | No | 9 | 5.5 | 7 | 6.5 | 1 | 5.9 | **17** | 4 | 4.7 | 1 | 2.3 | 11 | 2.8 | 2 | 3.9 | **18** | 2 | 8.0 | 2 | 4.5 | 11 | 6.0 | **15** |
| Total | |  | 164 | 100.0 | 108 | 100.0 | 17 | 100.0 | 289 | 86 | 100.0 | 43 | 100.0 | 395 | 100.0 | 51 | 100.0 | 574 | 25 | 100.0 | 44 | 100.0 | 182 | 100.0 | 251 |

Table 5. Convalescent anti-N seroconversion proportion by demographic characteristics, vaccination status, symptoms and comorbidities

|  |  | Anti-N Seroconversion | | | |  |  |
| --- | --- | --- | --- | --- | --- | --- | --- |
|  |  | Yes | | No | | Total | *p-value* |
|  |  | n | % | n | % | n |  |
|  |  | 1,065 | 95.5 | 50 | 4.5 | 1,115 |  |
| **Gender** | Female | 666 | 62.5 | 38 | 76.0 | 704 | *0.05* |
|  | Male | 399 | 37.5 | 12 | 24.0 | 411 |  |
| **Ethnicity** | White | 991 | 93.4 | 49 | 98.0 | 1040 | *0.19* |
|  | non-white | 70 | 6.6 | 1 | 2.0 | 71 |  |
| **Age** | 18-29 | 83 | 7.8 | 4 | 8.0 | 87 | *0.99* |
|  | 30-39 | 184 | 17.3 | 9 | 18.0 | 193 |  |
|  | 40-49 | 296 | 27.7 | 13 | 26.0 | 309 |  |
|  | 50-59 | 303 | 28.5 | 16 | 32.0 | 319 |  |
|  | 60-69 | 151 | 14.2 | 6 | 12.0 | 157 |  |
|  | 70+ | 48 | 4.5 | 2 | 4.0 | 50 |  |
| **Vaccination status** | unvaccinated | 260 | 24.4 | 15 | 30.0 | 275 | *0.25* |
|  | >28 days post D1 | 143 | 13.4 | 8 | 16.0 | 151 |  |
|  | >14 days post D2 | 442 | 41.5 | 14 | 28.0 | 456 |  |
|  | >14 days post D3+ | 220 | 20.7 | 13 | 26.0 | 233 |  |
| **Variant** | Alpha | 272 | 25.5 | 17 | 34.0 | 289 | *0.07* |
|  | Delta | 557 | 52.3 | 18 | 36.0 | 575 |  |
|  | Omicron | 236 | 22.2 | 15 | 30.0 | 251 |  |
| **Immunosuppressed** | Yes | 39 | 3.7 | 4 | 8.0 | 43 | *0.12* |
|  | No | 1,026 | 96.3 | 46 | 92.0 | 1,072 |  |
| **CEV** | Yes | 55 | 5.2 | 6 | 12.0 | 61 | *0.04* |
|  | No | 1,010 | 94.8 | 44 | 88.0 | 1,054 |  |
| **At risk** | Yes | 227 | 21.3 | 13 | 26.0 | 240 | *0.43* |
|  | No | 838 | 78.7 | 37 | 74.0 | 875 |  |
| **Comorbidity count** | 0 | 568 | 53.3 | 27 | 54.0 | 595 | *0.01* |
|  | 1 | 277 | 26.0 | 6 | 12.0 | 283 |  |
|  | 2 | 156 | 14.7 | 9 | 18.0 | 165 |  |
|  | 3+ | 64 | 6.0 | 8 | 16.0 | 72 |  |
| **Comorbidity** | Yes | 497 | 46.7 | 23 | 46.0 | 520 | *0.93* |
|  | No | 568 | 53.3 | 27 | 54.0 | 595 |  |
| **Symptoms** | Yes | 1,046 | 98.2 | 48 | 96.0 | 1,094 | *0.24* |
|  | No | 19 | 1.8 | 2 | 4.0 | 21 |  |
| **Severity** | Asymptomatic | 18 | 2.0 | 2 | 4.4 | 20 | *0.27* |
|  | Mild | 523 | 57.4 | 30 | 65.2 | 553 |  |
|  | Moderate | 341 | 37.4 | 14 | 30.4 | 355 |  |
|  | Severe | 29 | 3.2 | 0 | 0.0 | 29 |  |
| **Chronic respiratory disease** | Yes | 29 | 2.7 | 4 | 8.0 | 33 | *0.06* |
|  | No | 1,036 | 97.3 | 46 | 92.0 | 1,082 |  |
| **Chronic heart disease** | Yes | 42 | 3.9 | 5 | 10.0 | 47 | *0.04* |
|  | No | 1,023 | 96.1 | 45 | 90.0 | 1,068 |  |
| **Chronic kidney disease** | Yes | 5 | 0.5 | 0 | 0.0 | 5 | *0.80* |
|  | No | 1,060 | 99.5 | 50 | 100.0 | 1,110 |  |
| **Chronic liver disease** | Yes | 8 | 0.8 | 2 | 4.0 | 10 | *0.07* |
|  | No | 1,057 | 99.2 | 48 | 96.0 | 1,105 |  |
| **Diabetes** | Yes | 54 | 5.1 | 2 | 4.0 | 56 | *0.54* |
|  | No | 1,011 | 94.9 | 48 | 96.0 | 1,059 |  |
| **Chronic disease** | Yes | 126 | 11.8 | 11 | 22.0 | 137 | *0.03* |
|  | No | 939 | 88.2 | 39 | 78.0 | 978 |  |
| **BMI 40** | Yes | 42 | 3.9 | 4 | 8.0 | 46 | *0.15* |
|  | No | 1,023 | 96.1 | 46 | 92.0 | 1,069 |  |
| **Asthma** | Yes | 128 | 12.0 | 9 | 18.0 | 137 | *0.21* |
|  | No | 937 | 88.0 | 41 | 82.0 | 978 |  |
| **Immunosuppressed** | Yes | 39 | 3.7 | 4 | 8.0 | 43 | *0.12* |
|  | No | 1,026 | 96.3 | 46 | 92.0 | 1,072 |  |
| **Neurological disease** | Yes | 17 | 1.6 | 2 | 4.0 | 19 | *0.21* |
|  | No | 1,048 | 98.4 | 48 | 96.0 | 1,096 |  |
| **Cardiovascular disease** | Yes | 10 | 0.9 | 1 | 2.0 | 11 | *0.40* |
|  | No | 1,055 | 99.1 | 49 | 98.0 | 1,104 |  |
| **Hypertension** | Yes | 92 | 8.6 | 5 | 10.0 | 97 | *0.74* |
|  | No | 973 | 91.4 | 45 | 90.0 | 1,017 |  |
| **Endocrine** | Yes | 55 | 5.2 | 1 | 2.0 | 56 | *0.27* |
|  | No | 1,010 | 94.8 | 49 | 98.0 | 1,059 |  |
| **Mental health disease** | Yes | 38 | 3.6 | 1 | 2.0 | 39 | *0.47* |
|  | No | 1,027 | 96.4 | 49 | 98.0 | 1,076 |  |
| **Arthritis** | Yes | 32 | 3.0 | 2 | 4.0 | 34 | *0.46* |
|  | No | 1,033 | 97.0 | 48 | 96.0 | 1,081 |  |
| **Cancer** | Yes | 25 | 2.3 | 1 | 2.0 | 26 | *0.67* |
|  | No | 1,040 | 97.7 | 49 | 98.0 | 1,089 |  |
| **Other condition** | Yes | 196 | 18.4 | 9 | 18.0 | 205 | *0.56* |
|  | No | 869 | 81.6 | 41 | 82.0 | 910 |  |

Table 6. Multivariable linear regression investigating the convalescent anti-N antibody level by vaccination status for all variants and by variants - geometric mean (95% CI) and adjusted geometric mean ratio (95% CI) of the log of convalescent Roche N are presented

|  |  | | **All variants** | | **Alpha** | | | **Delta** | | | **Omicron** | | |
| --- | --- | --- | --- | --- | --- | --- | --- | --- | --- | --- | --- | --- | --- |
|  | N  1,214 | | Geometric mean  (95% CI) | Adjusted geometric mean ratio (95% CI) | N  304 | Geometric mean  (95% CI) | Adjusted geometric mean ratio (95% CI) | N  644 | Geometric mean  (95% CI) | Adjusted geometric mean ratio (95% CI) | N  262 | Geometric mean  (95% CI) | Adjusted geometric mean ratio (95% CI) |
| **Vaccination** |  | |  |  |  |  |  |  |  |  |  |  |  |
| Unvaccinated | 296 | | 24.6 (20.5-29.4) | 1 | 176 | 22.6 (17.7-28.8) | 1 | 96 | 30.3 (22.3-41.0) | 1 | 24 | 19.6 (11.5-33.4) | 1 |
| >28 days post D1 | 156 | | 11.8 (9.3-15.1) | 0.46 (0.34-0.60)* | 110 | 10.4 (7.7-14.0) | 0.40 (0.28-0.59)* | 46 | 16.1 (10.6-24.4) | 0.48 (0.29-0.79)* | 0 | N/A | N/A |
| >14 days post D2 | 511 | | 13.1 (11.6-14.8) | 0.41 (0.32-0.53)* | 18 | 10.5 (4.9-22.3) | 0.47 (0.22-1.00) | 447 | 13.8 (12.2-15.8) | 0.40 (0.28-0.56)* | 46 | 8.4 (5.4-13.1) | 0.46 (0.23-0.94)* |
| >14 days post D3+ | 251 | | 11.7 (9.9-13.9) | 0.48 (0.34-0.67)* | 0 | N/A | N/A | 55 | 12.9 (8.9-18.7) | 0.40 (0.25-0.65)* | 196 | 11.4 (9.4-13.9) | 0.62 (0.33-1.15) |
| **Variants** |  |  | |  |  |  |  |  |  |  |  |  |  |
| Alpha | 304 | 16.3 (13.5-19.6) | | 0.67 (0.52-0.87)* |  |  |  |  |  |  |  |  |  |
| Delta | 644 | 15.6 (14.0-17.5) | | 1 |  |  |  |  |  |  |  |  |  |
| Omicron | 266 | 11.4 (9.6-13.5) | | 0.74 (0.56-0.97)* |  |  |  |  |  |  |  |  |  |
| **Gender** |  |  | |  |  |  |  |  |  |  |  |  |  |
| Female | 745 | 13.5 (12.2-15.1) | | 1 | 187 | 13.7 (10.7-17.6) | 1 | 405 | 14.9 (13.0-17.1) | 1 | 153 | 10.4 (8.3-13.1) | 1 |
| Male | 469 | 16.8 (14.7-19.2) | | 1.21 (1.02-1.44)* | 117 | 21.6 (16.5-28.1) | 1.28 (0.87-1.87) | 239 | 17.0 (14.0-20.5) | 1.15 (0.91-1.44) | 113 | 12.8 (9.9-16.6) | 1.27 (0.88-1.83) |
| **Ethnicity** |  |  | |  |  |  |  |  |  |  | 0 | N/A | N/A |
| Other ethnic groups | 81 | 22.0 (15.9-30.4) | | 1.62 (1.16-2.25)* | 32 | 23.0 (12.7-41.4) | 1.63 (0.90-2.97) | 38 | 25.1 (16.6-38.1) | 1.67 (1.05-2.65)* | 11 | 12.1 (4.2-35.1) | 1.26 (0.52-3.03) |
| White | 1,133 | 14.3 (13.1-15.6) | | 1 | 272 | 15.7 (12.9-19.1) | 1 | 606 | 15.2 (13.5-17.0) | 1 | 255 | 11.3 (9.5-13.5) | 1 |
| **Age (year old)** |  |  | |  |  |  |  |  |  |  |  |  |  |
| 18-29 | 96 | 17.4 (13.2-23.1) | | 0.94 (0.67-1.31) | 28 | 19.0 (10.0-35.8) | 1.13 (0.58-2.21) | 50 | 20.7 (14.5-29.7) | 0.87 (0.54-1.38) | 18 | 9.4 (5.3-16.7) | 0.90 (0.41-1.97) |
| 30-39 | 204 | 11.7 (9.5-14.5) | | 0.74 (0.58-0.96)* | 53 | 9.4 (5.6-15.7) | 0.68 (0.39-1.20) | 102 | 14.4 (11.1-18.6) | 0.76 (0.54-1.07) | 49 | 9.7 (6.3-15.1) | 0.82 (0.45-1.50) |
| 40-49 | 326 | 15.3 (13.1-17.9) | | 1.08 (0.87-1.34) | 69 | 19.0 (13.3-27.2) | 1.25 (0.76-2.04) | 195 | 15.0 (12.3-18.3) | 1.01 (0.77-1.32) | 62 | 12.9 (9.2-18.1) | 1.14 (0.67-1.96) |
| 50-59 | 354 | 14.8 (12.6-17.4) | | 1 | 91 | 17.3 (12.2-24.6) | 1 | 212 | 14.8 (12.0-18.2) | 1 | 51 | 11.4 (7.8-16.7) | 1 |
| 60-69 | 173 | 17.9 (14.5-22.0) | | 1.33 (1.02-1.74)* | 47 | 18.5 (12.0-28.5) | 1.11 (0.63-1.94) | 64 | 22.9 (16.8-31.4) | 1.69 (1.14-2.51)* | 62 | 13.4 (9.4-19.2) | 1.11 (0.64-1.92) |
| 70+ | 61 | 11.1 (7.1-17.3) | | 0.86 (0.57-1.28) | 16 | 19.9 (8.1-48.7) | 1.59 (0.68-3.73) | 21 | 9.8 (4.4-22.0) | 0.77 (0.41-1.44) | 24 | 8.4 (4.1-17.4) | 0.60 (0.29-1.25) |
| **Comorbidity** |  |  | |  |  |  |  |  |  |  |  |  |  |
| No | 646 | 14.6 (13.0-16.3) | | 1 | 155 | 14.7 (11.3-19.2) | 1 | 384 | 15.8 (13.7-18.3) | 1 | 107 | 10.7 (8.3-13.8) | 1 |
| Yes | 568 | 14.9 (13.2-16.9) | | 1.08 (0.90-1.29) | 149 | 18.2 (14.0-23.6) | 1.00 (0.68-1.48) | 260 | 15.4 (12.9-18.3) | 1.09 (0.86-1.38) | 159 | 11.9 (9.4-14.9) | 1.09 (0.74-1.63) |
| **Immunosuppressed and CEV status** |  | |  |  |  |  |  |  |  |  |  |  |  |
| Neither | 1,119 | | 15.0 (13.7-16.3) | 1 | 281 | 15.7 (13.0-19.0) | 1 | 601 | 16.2 (14.5-18.1) | 1 | 237 | 11.6 (9.7-13.8) | 1 |
| Immunosuppressed only | 28 | | 8.8 (4.5-17.1) | 0.55 (0.32-0.95)* | 5 | 33.2 (9.3-118.8) | 2.13 (0.52-8.68) | 13 | 8.5 (3.3-22.1) | 0.45 (0.21-1.00) | 10 | 4.8 (1.2-18.4) | 0.39 (0.15-0.99)* |
| Immunosuppressed and CEV | 24 | | 7.5 (3.0-18.8) | 0.50 (0.28-0.91)* | 5 | 5.8 (0.1-307.7) | 0.40 (0.10-1.65) | 10 | 5.7 (1.1-28.7) | 0.38 (0.16-0.92)* | 9 | 11.7 (3.6-38.6) | 0.91 (0.35-2.41) |
| CEV only | 43 | | 20.1 (12.1-33.5) | 1.49 (0.95-2.35) | 13 | 42.0 (18.1-97.6) | 3.18 (1.26-7.98)* | 20 | 13.4 (5.9-30.3) | 0.92 (0.48-1.76) | 10 | 17.3 (5.3-56.2) | 1.77 (0.69-4.53) |
| **Time since event (days)** | 1,214 |  | | 1.02(1.01-1.03)* | 304 |  | 1.03 (1.01-1.05)* | 644 |  | 1.02 (1.00-1.03)* | 266 |  | 1.00 (0.97-1.03) |

* p<0.05

Interaction between vaccination status and variants was not statistically significant

All variants: Global LRT p-value for age: p=0.0053; Global LRT p value for immunosuppressed and CEV status: p=0.0038

Table 7.Ratio of convalescent and baseline anti-N antibody levels by vaccination status and variants

|  |  | All variants |  | Alpha |  | Delta |  | Omicron |
| --- | --- | --- | --- | --- | --- | --- | --- | --- |
|  | N  587 | Geometric mean  (95% CI) | N  65 | Geometric mean  (95% CI) | N  349 | Geometric mean  (95% CI) | N  173 | Geometric mean  (95% CI) |
| **Vaccination** |  |  |  |  |  |  |  |  |
| Unvaccinated | 103 | 254.8 (190.2-341.4) | 33 | 176.5 (102.3-304.3) | 50 | 399.6 (269.9-591.5) | 20 | 151.8 (77.3-298.1) |
| Vaccinated | 484 | 121.9 (107.5-138.2) | 32 | 98.6 (59.5-163.4) | 299 | 132.5 (112.6-155.8) | 153 | 108.2 (87.1-134.5) |

Table 8. Multivariable linear regression investigating the convalescent anti-S antibody level by vaccination status for all variants and by variants - geometric mean (95% CI) and adjusted geometric mean ratio (95% CI) of the log of convalescent Roche S are presented

|  | **All variants** | | | **Alpha** | | | **Delta** | | | **Omicron** | | | |
| --- | --- | --- | --- | --- | --- | --- | --- | --- | --- | --- | --- | --- | --- |
|  | N  1,214 | Geometric mean  (95% CI) | Adjusted geometric mean ratio (95% CI) | N  304 | Geometric mean  (95% CI) | Adjusted geometric mean ratio (95% CI) | N  644 | Geometric mean  (95% CI) | Adjusted geometric mean ratio (95% CI) | | N  266 | Geometric mean  (95% CI) | Adjusted geometric mean ratio (95% CI) |
| **Vaccination** |  |  |  |  |  |  |  |  |  | |  |  |  |
| Unvaccinated | 296 | 151.7 (114.1-201.6) | 0.01 (0.004-0.01)* | 178 | 225.4 (164.0-309.9) | 0.02 (0.01-0.06)* | 97 | 121.8 (76.8-193.2) | 0.01 (0.004-0.01)* | | 24 | 18.9 (3.5-103.2) | 0.001 (0.0003-0.002)* |
| AZ >28 days post D1 | 76 | 7,900.0 (6,085.0-10,256.4) | 0.68 (0.37-1.26) | 52 | 5,806.0 (4,389.6-7,679.5) | 0.39 (0.11-1.39) | 24 | 15,396.3 (9,445.4-25,096.3) | 0.67 (0.39-1.14) | | 0 | N/A | N/A |
| AZ >14 days post D2/D3 | 361 | 22,766.9 (20,685.2-25,058.1) | 1 | 9 | 11,739.0 (8,498.3-16,215.4) | 1 | 338 | 23,340.7 (21,095.6 -25,824.7) | 1 | | 18 | 19,769.3 (13,314.2-29,354.0) | 1 |
| PF/MD>28 days post D1 | 80 | 12,383.5 (9,200.1-16,668.3) | 0.96 (0.50-1.85) | 58 | 10,313.8 (7,072.7-15,040.0) | 0.81 (0.23-2.86) | 22 | 20,055.7 (13,542.8-29,700.7) | 1.00 (0.56-1.79) | | 0 | N/A | N/A |
| PF/MD>14 days post D2 | 151 | 33,129.3 (27,859.6-39,395.8) | 1.73 (1.26-2.38)* | 9 | 13,264.9 (6,667.2-26,391.6) | 1.20 (0.23-6.34) | 113 | 36,781.5 (29,978.7-45,127.9) | 1.77 (1.34-2.34)* | | 29 | 29,282.5 (20,641.4-41,541.0) | 1.27 (0.48-3.34) |
| PF/MD>14 days post D3+ | 250 | 35,947.5 (31,407.0-41,144.4) | 1.83 (1.19-2.80)* | 0 |  |  | 54 | 39,096.5 (28,524.8-53,586.4) | 1.87 (1.29-2.71)* | | 197 | 35,191.9 (30,308.7-40,862.0) | 1.70 (0.77-3.79) |
| **Variant** |  |  |  |  |  |  |  |  |  | |  |  |  |
| Alpha | 304 | 1,039.5 (779.8-1,385.6) | 0.51 (0.19-1.37) |  |  |  |  |  |  | |  |  |  |
| Delta | 644 | 11,774.0 (9,842.6-14,084.4) | 1 |  |  |  |  |  |  | |  |  |  |
| Omicron | 266 | 16,937.9 (12,313.3-23,299.4) | 0.94 (0.46-1.93) |  |  |  |  |  |  | |  |  |  |
| **Gender** |  |  |  |  |  |  |  |  |  | |  |  |  |
| Female | 745 | 6,753.4 (5,554.3-8,211.4) | 1 | 187 | 974.8 (660.1-1,439.4) | 1 | 405 | 11,087.9 (8,766.4-14,024.3) | 1 | | 153 | 19,362.7 (13,328.5-28,128.8) | 1 |
| Male | 469 | 7,256.1 (5,712.5-9,216.8) | 1.13 (0.95-1.35) | 117 | 1,152.0 (758.2-1,750.2) | 1.12 (0.72-1.73) | 239 | 13,034.9 (9,903.1-17,157.3) | 1.15 (0.93-1.42) | | 113 | 14,131.4 (8,070.9-24,742.6) | 1.05 (0.70-1.56) |
| **Ethnicity** |  |  |  |  |  |  |  |  |  | |  |  |  |
| Other | 81 | 5,269.0 (3,053.3-9,092.3) | 1.35 (0.96-1.90) | 32 | 1,502.8 (619.2-3,647.3) | 1.47 (0.74-2.92) | 38 | 7,942.7 (3,850.7-16,383.3) | 1.22 (0.80-1.86) | | 11 | 49,079.5 (26,396.2-91,255.5) | 1.68 (0.63-4.44) |
| White | 1,133 | 7,081.7 (6,050.3-8,288.9) | 1 | 272 | 995.4 (733.5-1,350.7) | 1 | 606 | 12,068.3 (10,027.6-14,524.2) | 1 | | 255 | 16,178.1 (11,621.8-22,520.8) | 1 |
| **Age** |  |  |  |  |  |  |  |  |  | |  |  |  |
| 18-29 | 96 | 1,908.9 (1,028.6-3,542.7) | 0.89 (0.63-1.27) | 28 | 698.2 (222.6-2,190.2) | 0.69 (0.32-1.48) | 50 | 1,617.8 (751.7-3,481.8) | 1.01 (0.65-1.57) | | 18 | 14,449.9 (2,738.3-76,252.3) | 0.86 (0.36-2.06) |
| 30-39 | 204 | 3,359.5 (2,178.1-5,181.8) | 0.70 (0.54-0.92)* | 53 | 799.4 (367.7-1,737.8) | 0.68 (0.36-1.29) | 102 | 3,682.7 (1,983.2-6,838.8) | 0.63 (0.46-0.87)* | | 49 | 13,111.5 (5,819.7-29,539.9) | 0.96 (0.49-1.89) |
| 40-49 | 326 | 7,568.9 (5,717.7- 10,019.4) | 0.75 (0.60-0.93)* | 69 | 752.1 (381.4-1,483.2) | 0.66 (0.38-1.16) | 195 | 13,828.1 (10,659.2-17,939.2) | 0.83 (0.65-1.07) | | 62 | 14,853.1 (7,261.3-30,382.1) | 0.59 (0.33-1.08) |
| 50-59 | 354 | 8,232.4 (6,319.7-10,724.1) | 1 | 91 | 1,006.3 (640.6-1,580.8) | 1 | 212 | 17,827.1 (13,575.6-23,410.0) | 1 | | 51 | 14,106.2 (6,138.3-32,417.0) | 1 |
| 60-69 | 173 | 13,866.8 (9930.8- 19,362.7) | 1.34 (1.02-1.77)* | 47 | 1,636.7 (863.2-3,103.1) | 1.54 (0.81-2.93) | 64 | 36,764.1 (28,201.7-47,926.2) | 1.41 (0.98-2.02) | | 62 | 25,607.5 (15,111.9-43,392.6) | 1.02 (0.56-1.87) |
| 70+ | 61 | 19,821.1 (12,882.4- 30,497.1) | 1.09 (0.72-1.66) | 16 | 6,371.1 (2,303.4-17,622.1) | 2.09 (0.79-5.53) | 21 | 39,874.2 (27,477.4-57,864.0) | 1.33 (0.74-2.37) | | 24 | 22,914.8 (10,946.9-47,966.9) | 0.54 (0.24-1.22) |
| **Comorbidity** |  |  |  |  |  |  |  |  |  | |  |  |  |
| No | 646 | 5,529.2 (4,455.2-6,862.2) | 1 | 155 | 785.8 (516.9-1,194.7) | 1 | 384 | 9,497.6 (7,460.6-12,090.8) | 1 | | 107 | 13,395.4 (7,516.1-23,873.4) | 1 |
| Yes | 568 | 8,996.1 (7,301.4-11,084.1) | 1.19(0.99-1.43) | 149 | 1,390.6 (939.9.-2,057.6) | 1.50 (0.97-2.33) | 260 | 16,171.0(12,451.4-21,001.8) | 1.14 (0.92-1.41) | | 159 | 19,835.3 (13,718.8-28,678.7) | 1.04 (0.67-1.60) |
| **Immunosuppressed and CEV status** |  |  |  |  |  |  |  |  |  | |  |  |  |
| Neither | 1,119 | 6,670.0 (5,695.9-7,810.6) | 1 | 281 | 1,018.7 (762.8-1,360.4) | 1 | 601 | 11,288.6 (9,347.7-13,632.4) | 1 | | 237 | 16,301.8 (11,534.7-23,039.0) | 1 |
| Immunosuppressed only | 28 | 7,429.3 (2,580.6-21,388.3) | 0.50 (0.29-0.89)* | 5 | 149.3 (3.29-6,782.8) | 0.29 (0.06-1.45) | 13 | 12,659.5 (3,306.7-48,466.8) | 0.53 (0.26-1.10) | | 10 | 26,212.8 (13,442.1-51,116.7) | 0.67 (0.24-1.86) |
| Immunosuppressed and CEV | 24 | 7,973.3 (2,080.2-30,560.9) | 0.66 (0.36-1.22) | 5 | 146.9 (0.28-78,197.0) | 0.24 (0.05-1.19) | 10 | 14,497.1 (5,747.3-36,567.8) | 0.85 (0.38-1.91) | | 9 | 37,744.2 (15,657.0-90,989.5) | 0.97 (0.33-2.82) |
| CEV only | 43 | 17,494.6 (9,300.0-32,909.9) | 1.07 (0.67-1.71) | 13 | 7,205.3 (2,423.7-21,420.6) | 0.94 (0.33-2.69) | 20 | 35,871.7 (22,383.9-57,486.7) | 1.20 (0.66-2.18) | | 10 | 13,183.5 (1,252.9-138,716.3) | 0.98 (0.34-2.77) |
| **Time since event (days)** | 1,214 |  | 1.01(1.00-1.02)* | 304 |  | 1.02 (1.00-1.05)* | 644 |  | 1.00 (0.99-1.01) | | 266 |  | 1.02 (0.98-1.05) |
| **Variant and vaccination** |  |  |  |  |  |  |  |  |  | |  |  |  |
| Alpha*Unvaccinated | 178 | 225.4 (164.0-309.9) | 3.11 (1.09-8.88)* |  |  |  |  |  |  | |  |  |  |
| Alpha*AZ >28 days post D1 | 52 | 5,806.0 (4,389.6-7,679.5) | 0.66 (0.20-2.23) |  |  |  |  |  |  | |  |  |  |
| Alpha*AZ >14 days post D2/D3 | 9 | 11,738.9 (8,498.3-16,215.4) | 1 |  |  |  |  |  |  | |  |  |  |
| Alpha*PF/MD>28 days post D1 | 58 | 10,313.8 (7,072.7-15,040.0) | 0.88 (0.26-2.99) |  |  |  |  |  |  | |  |  |  |
| Alpha*PF/MD>14 days post D2 | 9 | 13,264.9 (6,667.2-26,391.6) | 0.68 (0.17-2.77) |  |  |  |  |  |  | |  |  |  |
| Delta*Unvaccinated | 97 | 121.8 (76.8-193.2) | 1 |  |  |  |  |  |  | |  |  |  |
| Delta*AZ >28 days post D1 | 24 | 15,396.3 (9,445.4-25,096.3) | 1 |  |  |  |  |  |  | |  |  |  |
| Delta*AZ >14 days post D2/D3 | 338 | 23,340.7 (21,095.6 -25,824.7) | 1 |  |  |  |  |  |  | |  |  |  |
| Delta*PF/MD>28 days post D1 | 22 | 20,055.7 (13,542.8-29,700.7) | 1 |  |  |  |  |  |  | |  |  |  |
| Delta*PF/MD>14 days post D2 | 113 | 36,781.5 (29,978.7-45,127.9) | 1 |  |  |  |  |  |  | |  |  |  |
| Delta*PF/MD>14 days post D3+ | 54 | 39,096.5 (28,524.8-53,586.4) | 1 |  |  |  |  |  |  | |  |  |  |
| Omicron*Unvaccinated | 24 | 18.9 (3.5-103.2) | 0.15 (0.06-0.40)* |  |  |  |  |  |  | |  |  |  |
| Omicron*AZ >14 days post D2/D3 | 18 | 19,769.3 (13,314.2-29,354.0) | 1 |  |  |  |  |  |  | |  |  |  |
| Omcron*PF/MD>14 days post D2 | 29 | 29,282.5 (20,641.4-41,541.0) | 0.95 (0.37-2.44) |  |  |  |  |  |  | |  |  |  |
| Omicron*PF/MD>14 days post D3+ | 197 | 35,191.9 (30,308.7-40,862.0) | 0.86 (0.37-2.00) |  |  |  |  |  |  | |  |  |  |

* p<0.05 (All variants: Global LRT p-value for age: p=0.0003; Global LRT p value for immunosuppressed and CEV status: p=0.0572)

Table 9. Multivariable linear regression investigating the anti-S antibody levels in acute sera by vaccination type and doses - geometric mean (95% CI) and adjusted geometric mean ratio (95% CI) of the log of acute Roche S are presented

|  |  | **All variants** |  |
| --- | --- | --- | --- |
|  | N  108 | geometric mean  (95% CI) | Adjusted geometric mean ratio (95% CI) |
| **Vaccination** |  |  |  |
| AZ >28 days post D1 | 9 | 194.2 (33.6-1,123.1) | 0.22 (0.05-0.92)* |
| AZ >14 days post D2/D3 | 48 | 1,537.0 (933.8-2,529.8) | 1 |
| PF/MD>28 days post D1 | 7 | 1,287.0 (209.7-7,896.1) | 1.27 (0.30-5.43) |
| PF/MD>14 days post D2 | 20 | 3,357.0 (2,147.1-5,248.6) | 2.27 (0.94-5.51) |
| PF/MD>14 days post D3+ | 24 | 9,928.9 (6,860.6-14,369.4) | 9.94 (3.40-29.08)* |
| **Variant** |  |  |  |
| Alpha | 18 | 480.2 (171.6-1,343.6) | 0.43 (0.14-1.37) |
| Delta | 69 | 2,387.1 (1,594.1-3,574.6) | 1 |
| Omicron | 21 | 6,759.5 (4,108.5-11,121.1) | 0.42 (0.14-1.28) |
| **Gender** |  |  |  |
| Female | 67 | 2,209.8 (1,436.5-3,399.3) | 1 |
| Male | 41 | 2,282.6 (1,234.7-4,220.0) | 1.16 (0.62-2.17) |
| **Ethnicity** |  |  |  |
| White | 103 | 2,329.7 (1,635.7-3,318.3) | 1 |
| Other ethnic groups | 5 | 970.0 (61.6-15,263.1) | 0.72 (0.16-3.20) |
| **Age (year old)** |  |  |  |
| 18-39 | 28 | 2,291.7 (1,144.9- 4,587.0) | 0.93 (0.44-1.98) |
| 40-59 | 61 | 2,798.0 (1,781.7- 4,393.8) | 1 |
| 60+ | 19 | 1,052.9 (404.9- 2,737.9) | 0.47 (0.21-1.06) |
| **Comorbidity** |  |  |  |
| No | 58 | 2,442.4 (1,571.3-13,134.6) | 1 |
| Yes | 50 | 2,020.6 (1,144.5-3,567.4) | 1.10(0.59-2.04) |
| **Immunosuppressed and CEV status** |  |  |  |
| Neither | 102 | 2,316.0 (1,614.5- 3,322.3) | 1 |
| Immunosuppressed and/or CEV | 6 | 1,241.2 (209.1- 7,369.0) | 0.31 (0.08-1.20) |
| **Time since symptom onset (days)** | 108 |  | 1.49(1.06-2.09)* |

* p<0.05

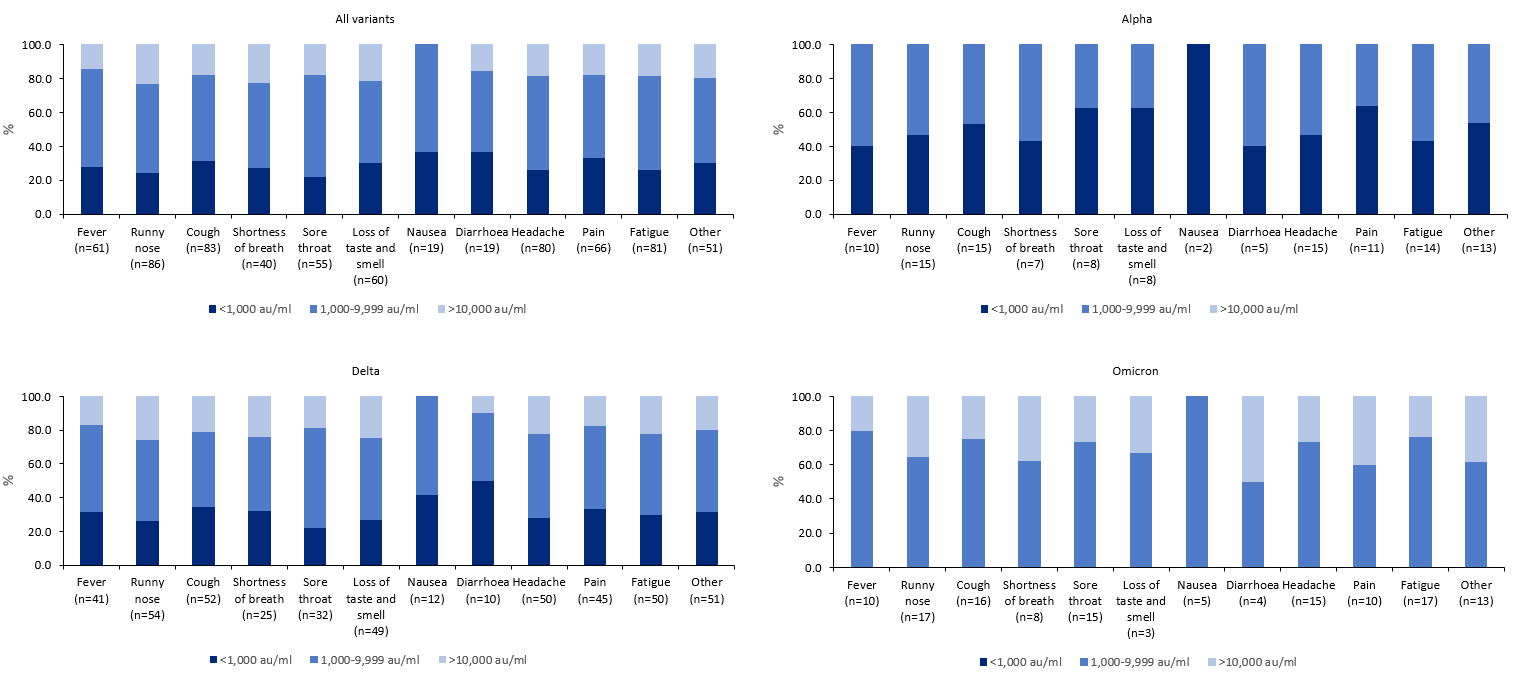

Figure 5. Anti-S antibody level in acute sera by symptoms type for all variants and by variants

**Supplementary materials for the methods section – description of the data sources**

**Data sources**

COVID-19 Testing Data

Community PCR testing (Pillar 2) was available to anyone with symptoms consistent with COVID-19 (high temperature, new continuous cough, or loss or change in sense of smell or taste), contacts of a confirmed case, care home staff and residents or individuals with positive lateral flow test (Figure 1, Supplementary materials). Pillar 2 cases were identified from the UKHSA Second Generation Surveillance System (SGSS) system or Unified Sample Database (USD) testing data. Information available includes demographic details, occupation information, PCR test result, date of symptom onset or cycle threshold (Ct) values. Testing data were linked to the National Immunisation Management System (NIMS) using a combination of National Health Service (NHS) number (a unique identifier for each person receiving medical care in the United Kingdom), date of birth, surname, first name and postcode and the variant data.

Vaccination data

Data on COVID-19 vaccination was extracted from NIMS, an electronic register of all COVID-19 vaccinations given in England (1). NIMS contains demographic information, dates of vaccination and vaccine manufacturer. NIMS contains some demographic information on the whole population of England who are registered with a general practitioner (GP) in England and is used to record all COVID-19 vaccinations.

Questionnaire data

Information on demographics details (gender, date of birth, ethnicity, address, number of people in household), vaccination history (COVID-19 vaccine: number of doses, brand, dates; date of last flu vaccine; vaccination against pneumococcus), COVID-19 illness (reason for COVID-19 test, source of infection, symptoms status, date of onset of symptoms, symptoms type, severity, date recovered, healthcare access, hospitalisation), medical history (clinically extremely vulnerable (CEV), chronic heart disease (CHD), diabetes, chronic kidney disease (CKD), chronic liver disease (CLD), chronic respiratory disease (CRD) excluding asthma, asthma requiring medication, malignancy, organ/bone marrow transplant, HIV/immunodeficiency, immunosuppression due to medication, seizure disorder, chronic neurological disease (CND), asplenia or dysfunction of the spleen, body mass index (BMI), pregnancy, history of previous infection), exposure risk factors (employment status, occupation, direct patient care, health and social care, number of days worked outside the home, travel) were collected at recruitment and 21 days post recruitment.

Serological testing

Samples were tested at the UKHSA national reference laboratory for the presence of antibodies to SARS-CoV-2 nucleoprotein (anti-N) and SARS-CoV-2 Spike Receptor Binding Domain (anti-S) using the “Elecsys anti-SARS-CoV-2 N” and the “Elecsys anti-SARS-CoV-2 S” electrochemiluminescence immunoassays respectively, on the Roche Cobas Pro e801 analyser. An anti-N antibody level of 1.0 cut-off index (COI) or greater was deemed positive and an anti-S antibody level of 0.8 arbitrary units per millilitre (AU/ml) or greater was deemed positive. An N assay positive result was considered indicative of prior SARS-CoV-2 infection, and an S assay positive result was considered indicative of prior SARS-CoV-2 infection or vaccination.

Oral fluid testing

Oral fluid samples were tested at the UKHSA national reference laboratory for SARS-COV-2 antibodies against an N gene and an S gene target. The collection and extraction of oral fluid from oracol swabs was undertaken as previously described (2, 3). Immunoglobulin capture assays for the detection of immunoglobulin G (IgG) were established for two targets: S1 and NP.

Virology data

Real-time reverse transcription PCR (RT-PCR) cycle threshold (Ct) values were used as a semiquantitative measure of SARS-CoV-2 viral load. Swabs were tested by a RT-PCR assay for both the ORF1ab and E gene regions of SARS-CoV-2. In a valid Orf1ab/E gene assay, any samples with cycle threshold (Ct) values ≤35 were considered positive, between 35 and 40 were considered indeterminate and >40 were considered negative. Positivity in both viral targets (ORF1ab and E gene) was required for the subject’s initial swab to be considered positive. For subsequent samples, single target positives could be accepted after assessment of data from previous samples.

Variant data

Variant status was confirmed by whole-genome sequencing of positive confirmatory PCR swabs for a subset of the samples, or sequencing results from original positive swab (where available). Variant results of all sequencing undertaken in England can be used to identify sequencing results for the original samples. If no sequencing result was available, S-gene target failure (SGTF) status from the original positive swab (if conducted using the TaqMan Assay) and date of test were used (SGTF is considered a proxy of Alpha for weeks 5 to 25; SGTF is considered a proxy of Omicron for week 48 onwards; S gene positive is considered a proxy for the Delta variant for dates from week 17 to 52). Individuals who could not be assigned a variant were excluded.

**References**

1.Tessier E, Edelstein M, Tsang C, Kirsebom F, Gower C, Campbell CNJ, et al. Monitoring the COVID-19 immunisation programme through a national immunisation Management system - England's experience. International journal of medical informatics. 2023;170:104974.

2.Hoschler K, Ijaz S, Andrews N, Ho S, Dicks S, Jegatheesan K, et al. SARS Antibody Testing in Children: Development of Oral Fluid Assays for IgG Measurements. Microbiology spectrum. 2022;10(1):e0078621.

3.Ijaz S, Dicks S, Jegatheesan K, Parker E, Katsanovskaja K, Vink E, et al. Mapping of SARS-CoV-2 IgM and IgG in gingival crevicular fluid: Antibody dynamics and linkage to severity of COVID-19 in hospital inpatients. The Journal of infection. 2022;85(2):152-60.
